# An interpretable omnigenic neural network architecture for the human genome

**DOI:** 10.64898/2026.07.28.26359187

**Authors:** Julius Upmeier zu Belzen, Lucas Arnoldt, Noah Hollmann, Luis Hermann, Khue M. Nguyen, Leonard Eckhoff, Leonhard Kohleick, Sedra Abou Ghaloun, Hannah Schmidt, Stefan Hegselmann, Fabian J. Theis, Thore Bürgel, Jakob Steinfeldt, Benjamin Wild, Roland Eils

## Abstract

Genetic prediction of complex phenotypes typically relies on additive linear models, which scale well but cannot capture non-additive effects or deeply integrate molecular and clinical data. Domain-specific neural networks have driven advances in images, text, and other modalities, but genome-scale neural networks remain challenging because genotypes are sparse and high-dimensional, effective sample sizes are limited, and generic architectures lack interpretability. Here, we introduce the omnigenic neural network, a biologically structured architecture inspired by the omnigenic model of complex traits. The model learns hierarchical representations of biological processes, accommodates multimodal inputs, supports transfer learning, and enables multitask prediction. Models trained in the UK Biobank and evaluated in the All of Us cohort for ischemic heart disease, type 2 diabetes, and schizophrenia outperformed published PGS Catalog and PRS-CSx scores. A multitask model trained across 36 cardiovascular endpoints further outperformed corresponding single-phenotype models and baselines. The architecture provides systems-level interpretability by quantifying the contributions of biological processes, which were consistent with established disease mechanisms. It also captures non-linear interactions between variants. Analysis of these interactions using Integrated Hessians revealed patterns concordant with previously reported epistatic associations. Together, these findings establish the omnigenic neural network as a flexible framework for interpretable, multimodal, and multitask genomic prediction.

## Introduction

Domain-specific neural network architectures have enabled revolutionary improvements in model performance for images^1^, text^2^, speech^3^, tabular data^4^, and other modalities, but remain underexplored for genomes so far. By exploiting fundamental properties of the data, such as translation invariance in images, or contextual dependencies in text, these architectures reduce parameter counts, improve training stability, provide interpretability, and scale substantially better than generic architectures. Gene-to-function models^5^ and language-model-inspired DNA-language models exist^6–8^, but are mostly limited to local applications, such as specific loci or genomic regions, rather than entire genomes. For genome-wide prediction, the current state of the art is to use Polygenic Scores (PGS)^9,10^, which are typically implemented as weighted sums and assume independent variant-effects. This assumption enables simple sharing and application to new datasets across existing frameworks^11^, use of summary statistics instead of individual-level genotypes and provides easy interpretation of individual variants effects. However, it ignores phenomena like epistasis^12^, or variant-covariate interactions^13^, which violate additivity. Neural networks however, are able to model such dependencies^14^. Aside from modelling non-additive aspects, PGS performance may be improved by increasing sample size, long-read sequencing, or multi-ancestry studies, all of which require substantial additional resources. Alternatively training models to predict multiple phenotypes simultaneously, especially pleiotropic ones, may improve performance, particularly for rare phenotypes^15,16^, but is difficult to directly integrate into PGS and GWAS workflows.

Notably, machine-learning approaches for improving PGS exist, but they tend to focus on improving phenotyping^17^ or covariates, for example by denoising them^18^, and still yield additive genotype-phenotype models. Neural networks to directly predict phenotypes based on genotypes have been developed^19,20^, but have not yet surpassed additive PGS^21^, or are narrowly tailored to specific phenotypes^20^, or require training data size exceeding even the largest biobanks^22^. Alternative machine learning approaches have been suggested as well, but are challenging to scale to genome-wide modeling, e.g. a recent work proposed XGBoost for obesity risk prediction, but was limited to 4000 variants^23^.

Engineering a neural network architecture that combines genome-wide scope, the capacity to model non-additive effects, and direct interpretability is genuinely difficult, for several key reasons. First, genotypes are extremely high-dimensional (768M variants reported in gnomAD³¹ version 4). Second, genotype sparsity and the low prevalence of many phenotypes in population cohorts necessitate large sample sizes, especially because high-effect variants tend to be rare³². Third, population-genetics software is optimised for variant-axis statistical analysis⁹,³³, whereas neural network training requires sample axis access. Fourth, the structure of a generic neural network is unrelated to the biology being modelled, so it remains a black box requiring post-hoc interpretability methods^24,25^, which are undesirable in clinical applications^26^. Fifth, populations of different genetic ancestry differ in allele frequencies, linkage disequilibrium (LD) patterns, and observed phenotype distributions. These differences may reflect both confounding and genuine genetic effects²⁷,²⁸, which is particularly relevant because over 80% of GWAS samples up to 2021 were of European ancestry²⁹. Ancestry-specific and multi-ancestry PGS methods aim to address this, with PRS-CSx³⁰ representing a state-of-the-art method.

Recent technical advances and increased data availability enable new approaches to address these challenges. Large cohorts like UK Biobank¹⁶,¹⁷ and All of Us¹⁸ provide genotype and health-record data for hundreds of thousands of individuals. Together with increasingly available high-performance GPUs, these resources make model training on individual-level data rather than summary statistics feasible, and provide the external validation needed to assess generalisation. Here, we explore an architecture that directly addresses these scalability and interpretability challenges: a neural network structured according to the Gene Ontology^27^ (GO) as an inductive bias. Specifically, the model can be considered a biologically informed sparse neural network, in which neurons are connected only when the corresponding biological entities are connected in the GO. In this architecture, each GO term is represented by a group of neurons whose inputs come only from the neurons of their child-terms, and outputs go only to their parent-term neurons. Thus, rather than parameterising all possible pairwise interactions, the model only parameterises interactions indicated by the GO. This type of model has seen some success in yeast-growth-rate modelling^28^, bulk transcriptomics^29^, single cell transcriptomics^30^ and disease-specific models^20^. However, applying it in a general population-genetics setting, would greatly increase the number of features from thousands for gene-level inputs, to hundreds of thousands or millions for variant-level inputs, which in turn requires more parameters and training samples.

To address these issues, we propose a neural network architecture for the human genome that scales to population cohorts, yields predictive PGS that are interpretable at multiple levels of abstraction and enables a range of novel analyses. It is a biologically informed sparse neural network based on the GO and extends this concept to the gene level by representing genes as nodes equivalent to GO-term nodes, and includes covariates at the gene level (Figure 1b). Consistent with Boyle et al.’s omnigenic model^31^, we represent gene effects through interconnected functional modules defined by the Gene Ontology, while learning the relationships among genes within each module during training. The specific implementation is inspired by the DCell model^28^ which was developed to predict yeast growth rates from binary gene knockout data. We substantially modify it, and extend it to the variant level and add covariate inclusion. We train our models on UK Biobank (UKB) data and externally evaluate them in the AoU cohort across a range of common disease phenotypes and compare them to published PGS^32^ and scores we constructed using PRS-CSx^33^, an established multi-ancestry PGS construction framework, together with published UKB summary statistics^34^.

**Figure 1.**
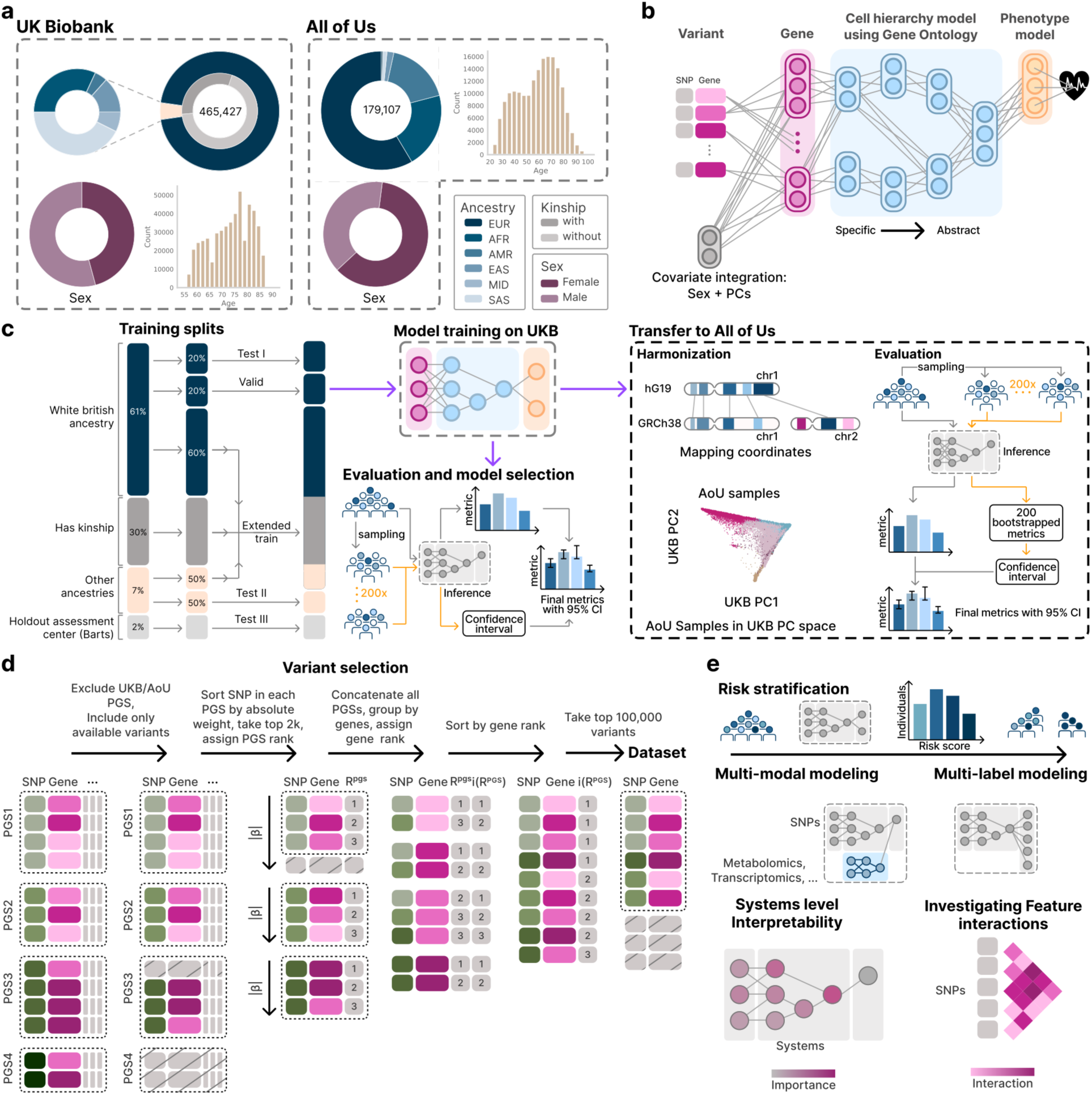
The OGM architecture enables phenotype prediction in AoU after training on UKB data. (**a**) The UKB cohort has mainly European ancestry, and covers an age range from about 55 to 85. The AoU population is much more diverse with substantial European, African and admixed American ancestries. It is younger, covering an age-range from 20 to 100, and is two-thirds female. (**b**) The OGM architecture has dosage inputs for genetic variants, as well as sex and genetic PCs as covariates into the first layer, representing individual genes. Gene representations are then fed to a GO-based cell-hierarchy-model that gets increasingly abstract, and at its root node is followed by a phenotype model. This can include further covariates and provides scores for a given phenotype. (**c**) The UKB data is split to ensure separation of training and testing data, control for different ancestries and flexibility for evaluation schemes. Specifically, a white British test and validation dataset without individuals with kinship are kept for model selection and evaluation. Similarly, a non-white British test set, and a set comprising a single assessment centre (Barts) are kept outside of the training data. The rest of the data is used for training. Metrics are computed with bootstrapping to obtain 95 % confidence intervals. To transfer models to the AoU population, the variant sets available are intersected, coordinates lifted over to hg38, and UKB eigenvalues are used to project AoU samples into the UKB PC space that is used for covariates. (**d**) 100,000 variants are selected to train models, based on publicly available PGS betas and gene assignments (see Methods). (**e**) A GO importance measure can be computed for each node in each model by fitting a logistic regression on the activation space to predict the phenotype in question and measuring its AUROC. The Integrated Gradients method can be used to infer the importance of individual variants for the model prediction. Integrated Hessians are used to determine how one single nucleotide polymorphism (SNP) influences another SNPs importance.

## Results

We introduce the OGM, named after the omnigenic model^31^, as a flexible, interpretable, domain-specific architecture for phenotype prediction from the human genome (Figure 1b). Specifically, dosage information for each variant is first passed to a gene layer, in which each gene is represented by 2 – 10 neurons. The resulting gene-level activations are passed to the GO model which is structured according to the biological process GO^27^. Each GO term is likewise represented by 2 – 10 neurons. Each gene’s activations are passed to GO terms with which it is annotated, and activations of GO terms are passed to their parent terms. Because the GO is a rooted directed acyclic graph, all information is ultimately aggregated at the root term of the ontology. A small single-layer phenotype model is then used to map this genotype representation to the phenotype for which the model is trained. The OGM architecture uses genetic sex to take sex-specific effects and confounding into account^13^. Similarly, it includes genetic principal components (PCs) to represent genetic ancestries and integrate them continuously rather than relying on discrete and potentially imperfect ancestry-boundaries^35^. We include these covariates directly in the gene layer, so that variant effects are always modelled in this context.

### Training, evaluation, application and interpretability

To assess whether OGM models generalise across ancestry groups and transfer to an independent cohort under realistic conditions, we trained the models in the UKB and evaluated them both internally in the UKB and externally in the All of Us (AoU) cohort (Figure 1a). Models were trained using data from all ancestries (Figure 1c). Internal evaluations in the UKB were performed separate for European and non-European ancestries ensuring that no related individuals were shared between training and evaluation sets. Because AoU has a more diverse ancestry distribution (Figure 1a), and none of its data was used for training, we were able to perform the external evaluation separately across continental-scale ancestry groups provided by AoU. These are based on the Phenotype Genotype Reference Map and include African ancestry (AFR), Latino/ admixed American ancestry (AMR), East Asian ancestry (EAS), European ancestry (EUR), and South Asian ancestry (SAS). To ensure that covariates were represented consistently, we projected the AoU genotypes into the UKB PC space (Figure 1c). We confirmed that this projected space substantially overlapped with the AoU-derived PC space and retained resolution of AoU ancestry groups (Supplementary Figure 1). Each model used an input set of 100,000 variants. This number represents a compromise between computational feasibility, including storage requirements, model size, and training speed on the one hand, and sufficiently broad coverage of the genome on the other (Figure 1d). Variants were selected based on their weights in published PGS that were not derived from UKB or AoU, while balancing the number of variants per gene (Methods). The resulting architecture supports single-label and multilabel risk prediction, multimodal modelling, and interpretability analyses at the systems, variant, and variant-interaction levels (Figure 1e).

### OGM-based polygenic scores

To evaluate whether the OGM architecture can be used as a practical PGS-like predictor across cohorts and ancestries, we trained OGM models for ischemic heart disease (IHD), type 2 diabetes (T2D), and schizophrenia (Figure 2a-c). These are relatively common phenotypes often used in PGS development. The models were trained in the UKB with sex and genetic PCs 1-3 as covariates and externally validated in the AoU dataset relative to published and custom PGS baselines including the same covariates.

**Figure 2.**
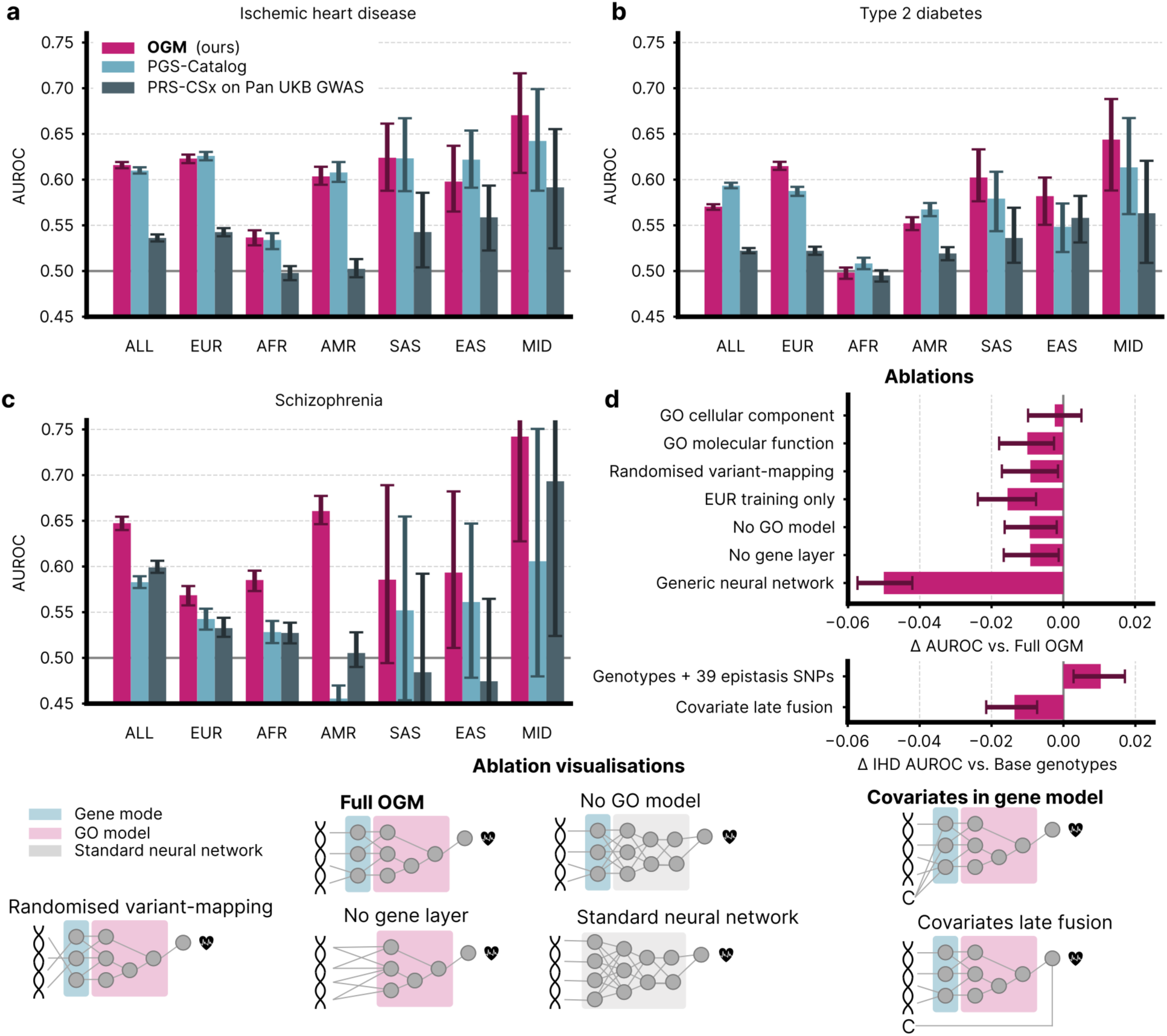
Performance of OGM-based and alternative PGS with sex and genetic PCs 1-3 as covariates. UKB performance for OGM-based PGS is measured in held-out, unrelated subpopulations. Models with covariates of non-UKB PGS are fitted on the UKB training split used for the OGM. AUROC for binary prediction of (**a**) ischemic heart disease (**b**) type 2 diabetes, and (**c**) schizophrenia (PGS000013, PGS004082, PGS000136, respectively). (**d**) Ablations of the OGM architecture. Models are for IHD without covariates in the upper plot, with covariates in the lower plot. We test the utility of the prior knowledge in the OGM by randomising the variant-to-gene mapping, replacing the GO-model with a standard neural network, removing the gene layer, and a comparison against a standard neural network. Additionally, we replace the biological process ontology with the cellular component and molecular function ontologies, and train on European-only datasets with and without individuals with kinship. Finally, we compare models trained with additional epistatic variants, and a late covariate fusion variant of the model with a standard OGM with covariates. 95%-Confidence intervals are obtained by bootstrapping the AUROC 200 times.

The OGM models reached AUROC scores of 0.623 for IHD, 0.615 for T2D and 0.569 for schizophrenia prediction in the EUR ancestry group. Across all ancestry groups, they outperformed the PRS-CSx baselines that are constructed from the PanUKBB summary statistics, which were based on the entire UKB population without hold-out data for internal evaluation (see Methods). To compare the OGM with established externally developed scores, we next evaluated its performance against published PGS across ancestry groups. For IHD, the OGM models outperformed state of the art published PGS^10^ by 0.006 in the entire population, but matched its performance within confidence intervals for all ancestry groups individually. For T2D, the OGM scores had higher performance in the EUR and EAS populations, and matched the PGS baseline^36^ in the other groups. For schizophrenia, the OGM models consistently outperformed the PGS baseline^37^ in all ancestry groups, except in the SAS, EAS, and MID ancestry groups, where performance estimation is exceedingly noisy.

To determine which architectural components contributed to OGM performance, we performed extensive ablation experiments using IHD phenotype prediction (Figure 2d). These experiments assessed the benefit of the OGM relative to standard neural networks, the contributions of its gene model and GO-model, and the effect of covariate integration in the gene layer. We further found that using the biological process GO leads to better performance than using the other GO namespaces or a randomised structure (further details in Methods).

### Model interpretability

To examine which biological processes and genetic variants drive OGM predictions, we assessed model interpretability at three levels. First, GO importance quantifies how predictive the learned representation of each GO term is of the phenotype (Figure 3a). For each term this is computed as a linear probe by fitting a logistic regression on its activation space to predict the phenotype and measuring the resulting AUROC. Second, via Integrated Gradients (IG)^38^ we quantified the effect of each variant on the model’s predictions. Third Integrated Hessians^38^ (IntH) quantify how one variant modifies the effect of another variant, which is the non-additive contribution of the variant pair.

**Figure 3.**
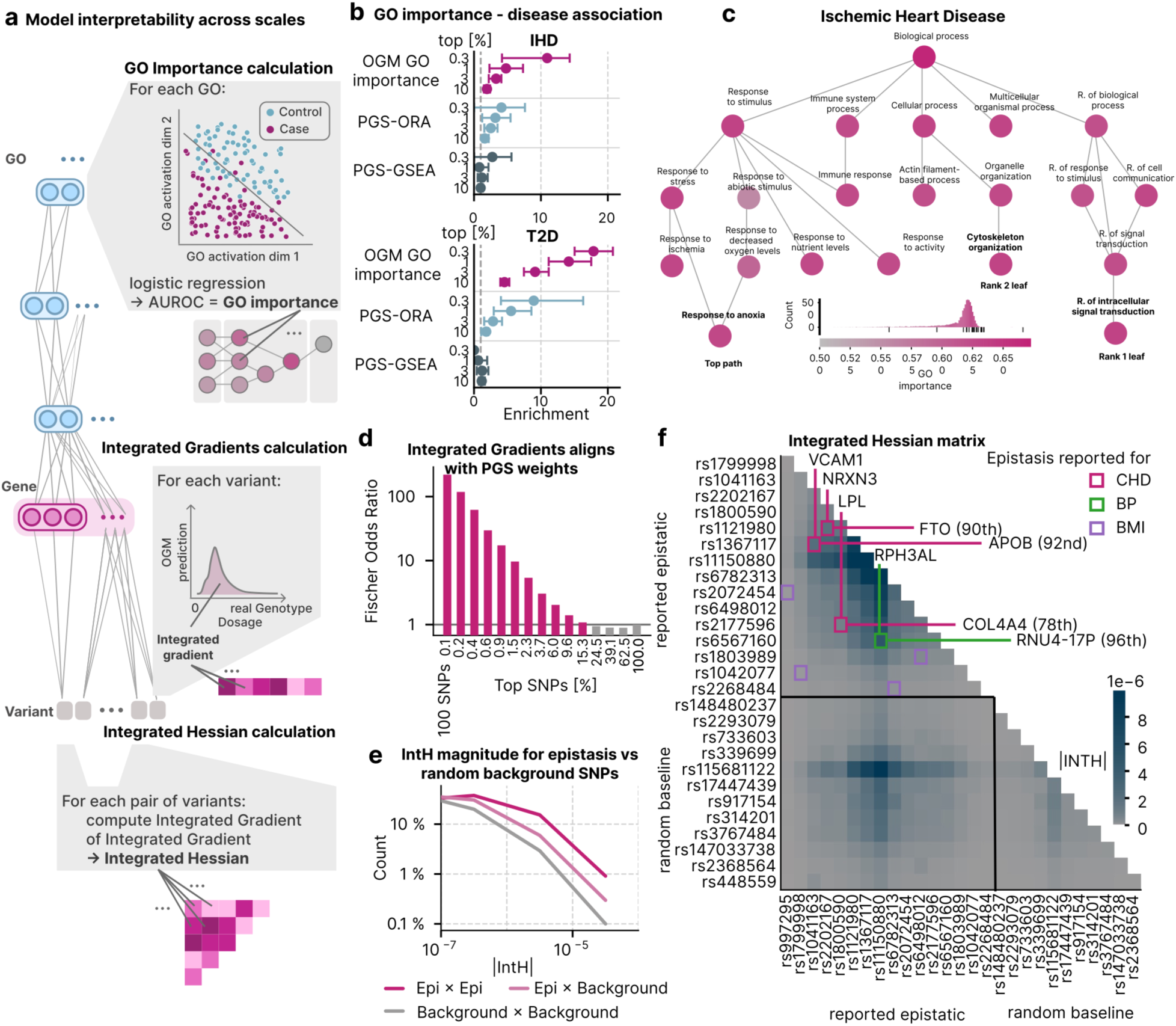
The OGM architecture enables interpretability across scales and recapitulates disease biology and epistasis. (**a**) Overview of interpretability mechanisms. At the systems-level we compute the GO importance to quantify how predictive each GO terms representation is for the predicted phenotype. Integrated Gradients quantify the contribution of each individual variant to the model’s prediction; Integrated Hessians extend this to quantify how one variant’s contribution is influenced by another, capturing pairwise variant-interactions learned by the model. (**b**) High OGM GO importances are enriched for terms associated with the predicted phenotype. The baselines are an overrepresentation analysis (ORA), and a gene-set enrichment analysis (GSEA) using weights of PGS000013 for coronary artery disease. 95%-Confidence intervals are obtained by bootstrapping the enrichment 200 times. (**c**) GO importances for a single-label IHD model are shown for the top paths, and top two leaves, including all their parent terms with no manual exclusions. Nodes are coloured by GO importance; the colour scale includes a histogram of all GO importances in the model with black markers indicating the values for the nodes shown in the figure. (**d**) Fisher odds ratio of overlap between the top-k SNPs ranked by |IG| and by |PGS000013| weight, across logarithmic thresholds from 0.1 % (100 SNPs) to 100 %. Bars coloured by OR>1. (**e**) Histograms of IntH upper 20 % of IntH values for Epi×Epi, Epi×other, and other×other combinations with logarithmic y-scale. (**f**) IntH heatmap for literature-reported epistatic pairs and random baseline variants. Axis labels show gene name and rsID; coloured squares mark known pairs.

We next investigated whether the processes prioritised by the models aligned with established disease biology and may be useful for hypothesis generation. To this end, we computed GO importance values for the IHD and T2D models (Supplementary Figure 2). In the IHD model, the top-down path with the highest GO importance values lead to the term *Response to anoxia* (Figure 3c), in line with the fundamental role oxygen-supply levels play in ischemic heart disease^39^. Other high-ranking nodes diverging from the top path included *immune response* and *response to nutrient levels*, which likely reflect the immune systems role in atherosclerotic lesion development^40^, and metabolic basis of IHD^41^. The two highest-ranked leaf nodes were *Regulation of intracellular signal transduction,* a high-level term whose high rank likely reflects its generality rather than IHD-specific biology, and *Cytoskeleton organisation,* which serves signalling roles in vascular mechanosensing^42^. In the T2D model (Supplementary Figure 2), the top path and top leaf terms contain a variety of signaling terms like ‘*Cell-surface receptor signaling pathway’*, ‘*Regulation of signal transduction*’, ‘*Hormone-mediated signaling pathway’*, as well as ‘*Regulation of cellular biosynthetic process’*, which leads to ‘*Regulation of metabolic process’*, all in line with the central role of insulin signaling biology underlying T2D^43^. To test this concordance systematically, we checked how well GO importances predicted whether the respective GO term is associated with the disease in published mappings^44,45^ (see Methods). We find that terms with high GO importance enriched significantly stronger for disease-associated GO terms than the overrepresentation analysis (ORA)-based and gene set enrichment analysis (GSEA)-based baselines (see Methods), for both IHD and T2D. To better understand model behaviour at the variant input level, we compared the IG^38,46^ values of the IHD model with the variant weights of a published polygenic risk score for coronary artery disease, which represents a substantial subset of IHD (PGS000013^10^). Variants with the highest IG values and PGS weights showed strong overlap, with a Fisher’s odds ratios of 220 for the top 100 (0.1%) of SNPs. This corresponds to an overlap of 20 of the top 100 SNPs. Fischers odds ratios are above 1 across the top 10 % of IG values (Figure 3d). This confirms that the model recovers established strong-effect variants, whereas variants of moderate importance may be prioritised differently, particularly if they are in linkage disequilibrium. Locus-level inspection of six representative genes contributing to this overlap is shown in 3 and confirms this concordance at variant-resolution.

### Integrated Hessians recover non-additive variant-interactions enriched at known epistatic loci

Unlike additive PGS, the OGM architecture can model non-additive effects like epistasis. We thus sought to identify variant pairs with these interactions and compare them to previously reported epistatic interactions. To this end, we computed IntH values^38^ for an epistasis variant set of 39 variants including 14 literature-curated epistatic pairs associated with IHD (5 pairs), blood pressure (2 pairs) and BMI (7 pairs), and a randomly selected background set of 100 variants in the UKB data (see Methods). Including the epistasis variants in the training datasets improved models AUROC by 0.01 relative to a model trained without them (Figure 2d), supporting their functional relevance to the model’s learned risk representation. IntH values were significantly higher among epistasis variants (Epi×Epi) than between epistasis and background variants (Epi × background, one-sided Mann-Whitney U test Bonferroni-corrected p=5e-33) or among the background variants (background × background, p=2e-79). Similarly, the Epi× background IntH values were higher than background × background (p=2e-30, Figure 3e). The ten highest IntH values all involved either RPH3AL, a regulator of insulin secretion^47^, or APOB^48^, a lipid carrier strongly linked to lipid metabolism and associated cardiovascular diseases, and targeted by a variety of lipid-lowering therapies (Supplementary Table 1).

To investigate the concordance of individual epistatic pairs with high-IntH pairs, we examined the IntH values for 16 epistasis variants and 11 random variants in detail (Figure 3f). The IHD-associated pairs rank in the 92^nd^, 90^th^, and 78^th^ percentiles. The pairs reported for the related phenotypes are in the 96^th^ percentile for blood pressure, between the 10^th^ and 66^th^ percentiles for BMI. Although this is limited by the small number of high-confidence reports of epistatic interactions and the very high computational cost of IntH calculations, the results support non-additive behaviour in the OGM-based models and show a substantial concordance between IntH values and reported epistasis for the same phenotype. This overlap was weaker for interactions reported for related phenotypes; nonetheless, variants characterised as epistatic for these phenotypes still tended to have elevated IntH values. Notably, the variants analysed here are distributed across genes and chromosomes, preventing the joint-tagging effect described by Kelemen et al.^21^ from distorting these interactions through linkage disequilibrium patterns.

### Multilabel predictions

Multilabel modelling may improve both computational efficiency and predictive performance when related phenotypes share underlying biological mechanisms. By predicting multiple labels simultaneously, machine-learning models can reuse shared representations and benefit from a richer training signal than models trained separately for each label^49^. Similar benefits have also been shown in population genetics^15^.

To test whether the OGMs can exploit shared genetic signals across related cardiovascular phenotypes, we trained a single model to predict all cardiovascular phecodes with more than 1,000 cases in the UKB. Cardiovascular diseases provide a suitable test case because they are widely studied in population genetics^10,50,51^, vary substantially in prevalence, and share some underlying genetics. In the EUR ancestry, our multilabel model outperformed the published PGS baseline for 71 % of phenotypes (one-sided Wilcoxon rank-sum test Holm-adjusted p=0.003), the PRS-CSx baseline for 94 % of phenotypes (p=0.0005), and single-label OGM models for 77 % of phenotypes (p=7e-5, Figure 4a, f). These improvements were observed across two orders of magnitude of phenotype incidence (Figure 4b), four orders of magnitude for the sample size of the GWAS used to construct the PGS baselines (Figure 4c), and five orders of magnitude for the number of variants included in the baseline PGS (Figure 4d). For the most common phenotypes like hypertension, performance differences could also be resolved in the smaller ancestry groups, where the multilabel model consistently outperformed the three baselines (Figure 4a, Supplementary Figure 4). Because performance estimates for less common phenotypes are less precise, we additionally assessed the proportion of phenotypes for which the multilabel model outperformed each baseline (Figure 4f). The multilabel model outperformed PRS-CSx with p < 0.002 in all but the AFR ancestry group (One-sided Wilcoxon test, Holm-adjusted p). Similarly, it had a higher AUROC than the PGSCatalog baselines across the EUR, AMR, SAS and MID ancestry groups with p < 0.05. Finally, it had significantly higher AUROC values than the single-label OGM models in all but the AFR population (adjusted p < 0.05). These results further indicate that the multilabel OGM model transfers more effectively across ancestries than the baselines. Performance in the AFR population was near random, and the smaller sample sizes of the MID and EAS populations lead to much noisier performance estimates (Figure 3g, Supplementary Figure 4). Notably, we observed the highest performance of the multilabel OGM for IHD prediction with an AUROC of 0.65, substantially above the single-label IHD models CI which only reached an AUROC of 0.62 (Figure 2a). This result illustrates the performance benefit of the multilabel model for one of the most common phenotypes. The improvement was even more pronounced for cardiomyopathy, for which the single-label OGM fails to achieve predictive performance, whereas the multilabel model reaches an AUROC of 0.62. Cardiomyopathy was approximately 10x less common (Figure 4b), making model fitting in population cohorts considerably more difficult and typically necessitating endpoint-specific datasets. Juergens *et al* used a total of six disease-specific and biobank datasets, including the UKB, to construct PGS004951. Nevertheless, this PGS was outperformed by the OGM trained using the UKB data alone, illustrating the capacity of multilabel models to utilize available training data much more effectively.

**Figure 4.**
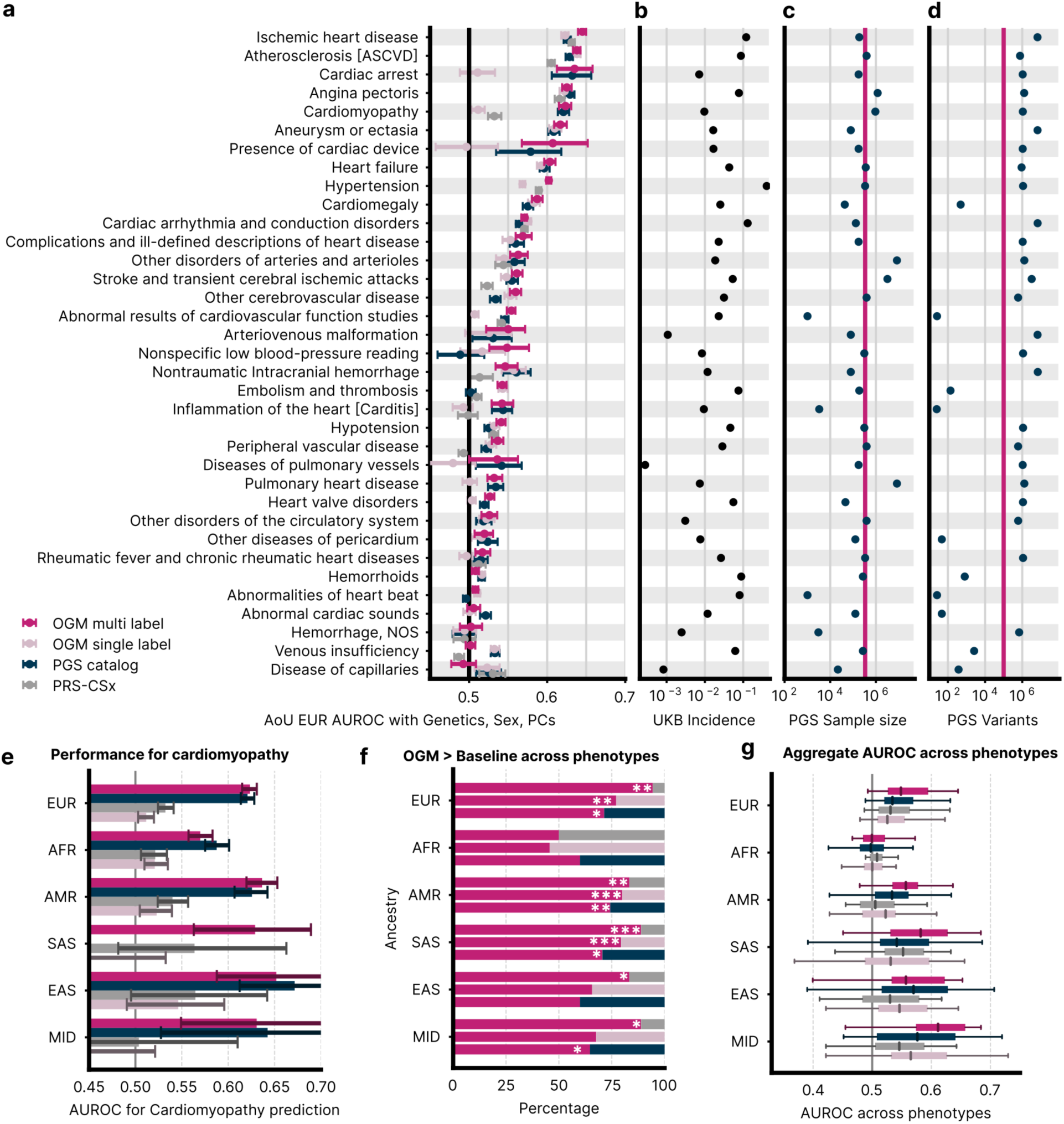
Multilabel performance across cardiovascular phenotypes. (**a**) All AUROC values are based on genotype, sex and genetic PCs. The multilabel and single-label OGMs are trained in the same set up on UKB data. PRS-CSx models use PanUKB GWAS. PGS models including covariates are fitted as logistic regressions on the UKB training data used for the OGMs and transferred to AoU. Confidence intervals are 95 percent, based on bootstrapping. (**b**) The incidence is computed on the UKB data. (**c**) The sample size and (**d**) number of variants are obtained from the PGS metadata, and the OGM training data. Confidence intervals are obtained by bootstrapping the AUROC 200 times. (**e**) Per ancestry performance of the multilabel model and its baselines for the prediction of cardiomyopathy. (**f**) Percentage of phenotypes for which the multilabel model outperforms the baselines per ancestry. * indicates p < 0.05, ** indicates p < 0.01, *** indicates p < 0.001 adjusted for multiple comparisons using the Holm-Bonferroni method. (**g**) Aggregate performance across ancestries. Boxplots indicate the median and interquartile range, whiskers indicate the most extreme values.

To investigate which biological processes contributed to the IHD predictions of the multilabel model, we examined GO-term importance scores for the IHD label in this model (Supplementary Figure 5). Highly important terms in the top path and two highest-ranked leaves (see Methods) included processes related to hypoxia, a central feature of IHD^52^, RNA-metabolism which have important roles in heart development, physiology, and pathology^53^, and immune response processes, which are targeted by anti-inflammatory treatment to reduce cardiovascular events in individuals with coronary heart disease^54^.

### Models based on genotypes and metabolites

In its hierarchical structure, the OGM learns rich representations across multiple levels of abstraction that can be used for model adaptations. To show this in practice, we sought to integrate metabolomic data with genetic data in the OGM architecture and demonstrate better prediction and interpretability for disease phenotypes. Specifically, we extended the OGM architecture with the ChEBI^55,56^ ontology, providing metabolite information alongside the GO-hierarchy (Figure 5a). We compared this model with single-modality models using genetics or metabolomics alone, as well as with a generic neural-network baseline that simply concatenated the metabolomic and genetic inputs. We trained all models on the UKB data to predict cardiovascular phecodes over a 10-year period following the metabolomics assessment (Figure 5b, see methods). To evaluate these models for incident risk prediction, we excluded individuals with diagnoses prior to the metabolomics assessment, separately for each phenotype. This notably removes the early-onset cases typically best predicted by genotype-based models.

**Figure 5.**
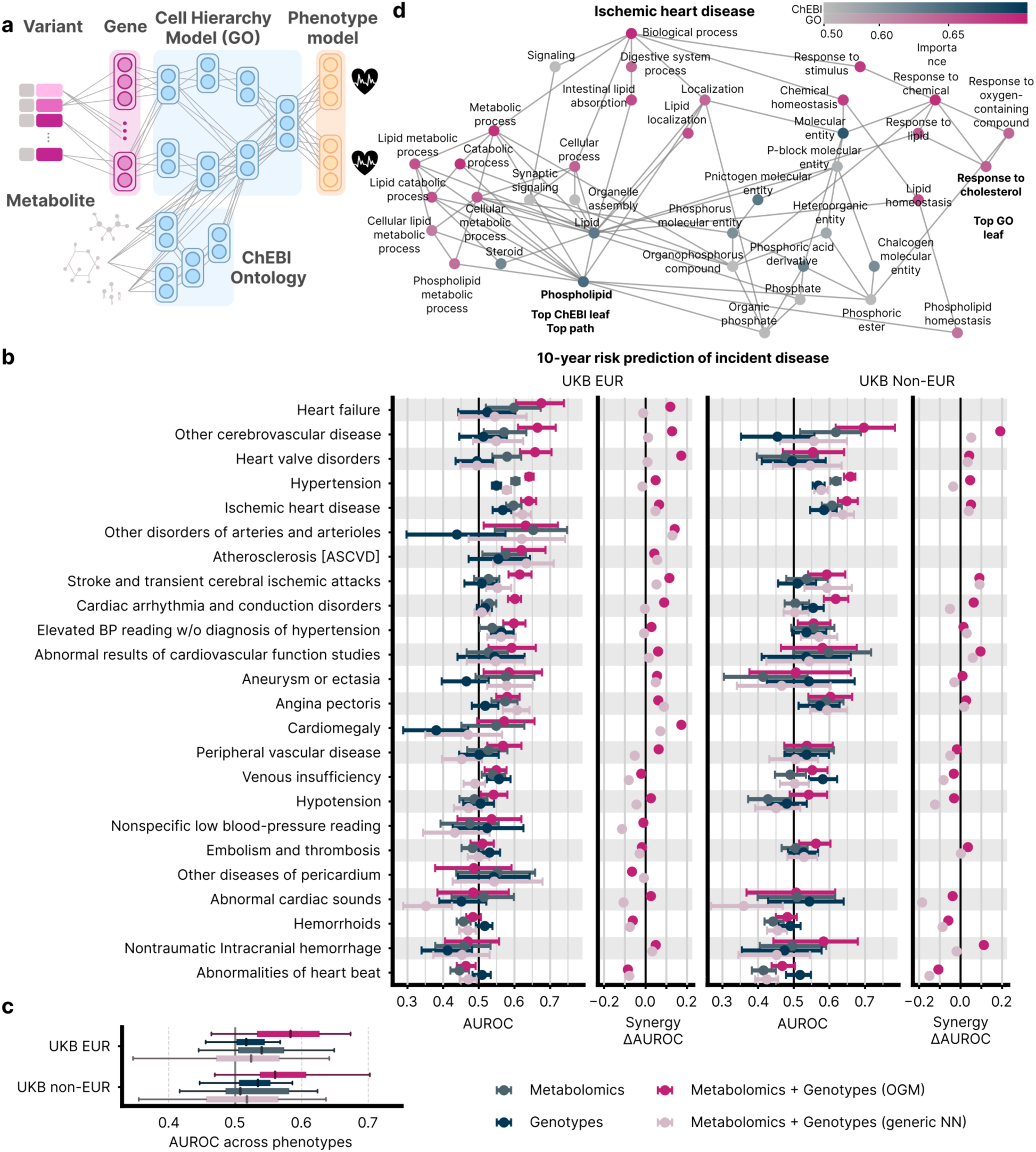
The OGM architecture incorporates interpretable metabolomic information for enhanced disease prediction. (**a**) The ChEBI ontology can be fused to the GO via a term-to-term mapping. Its inputs are the NMR metabolomics data provided by the UKB. (**b**) The full models using metabolomic and genetic data, only metabolomic data, only genetic data, and both in a fully-connected network (FC), are trained in the same setting for 10-year risk prediction. The model performance is evaluated as the AUROC over 10 years after the metabolomics assessment. For each phenotype, individuals with occurrences prior to that assessment are excluded from the evaluation. Phenotypes with fewer than 10 test cases are dropped from this figure. Confidence intervals are obtained by bootstrapping the AUROC 200 times. (**c**) Aggregate performance across ancestries. Boxplots indicate the median and interquartile range, whiskers indicate the most extreme values. (**d**) GO importances for the heart failure label are shown for the top path and the top leaf that is a ChEBI term, ‘*cholestanoid*’, including all their parent terms with manual exclusions. ChEBI terms have a blue colourscale. Nodes are coloured by GO importance; the colourscale includes a histogram of all GO importances in the model with black markers indicating the values for the nodes shown in the figure.

The combined OGM model outperformed the generic model for 83% of phenotypes in the EUR test set and 94% of phenotypes in the non-EUR test set (Figure 5b, c, Wilcoxon rank-sum test p < 0.001 for both). These results support the utility of the OGM architecture for model adaptation and further highlight its advantages in cross-ancestry settings. As expected, the combined model also consistently outperformed the genotype-only and metabolomics-only models in the EUR population for 87% and 79% of phenotypes (p < 0.001 for both), and in the non-EUR population for 83% and 72% of phenotypes, respectively (p < 0.001 and p = 0.01).

The combined model achieved its highest performance for heart failure prediction in the EUR ancestry with an AUROC of 0.67, compared with 0.59 for the metabolomics-only model and 0.53 for the genetics-only model. This AUROC improvement is substantially higher than those from earlier studies that integrate metabolomics and genetics data for disease risk prediction in the UKB^58,59,60^. More generally, phenotypes that were predicted well in the EUR population, are also tended to be predicted well in the non-EUR population: among the five best-predicted EUR phenotypes that were available in both groups, four were also among the five best-predicted phenotypes in the non-EUR population.

The additional ChEBI hierarchy allows us to extend our GO importance to chemical classes and their links into the GO graph (Figure 5d). GO and ChEBI importance scores for IHD (Figure 5d, pink and blue, respectively) identified the ChEBI term *Phospholipid* as the top leaf and the end of the top path, leading through *Lipid metabolic process*, *Intestinal lipid absorption*, and *Lipid homeostasis.* The top GO leaf node is *response to cholesterol*, the third highest ranked leaf overall. Notably, among the parent terms of *Lipid*, are terms like *Organelle assembly,* and *Synaptic signalling*, that are directly connected to the highly important *Lipid* node but have near-random GO importance themselves. Additionally, the *Signaling* term shows very low GO-importance as well, a marked contrast to the genotype-only GO importances that all show high importance for signaling terms (Figure 3c, Supplementary Figures 3, 5). Together, these findings suggest that after adding metabolomics data, the model’s importance shifts toward lipid-metabolism processes, consistent with the well-established role of lipid metabolism in IHD^57^.

To assess these effects more systematically, we compared GO-importance rankings between the combined model and a genotype-only multilabel model across their shared GO-term universe (Methods). The largest increase following the inclusion of metabolomic data was observed for *Biphenyl catabolic process*, for which GO importance increased from a near-random AUROC (0.473) in the genotype-only model to 0.629 in the combined model. This was followed by *Regulation of NAD(P)H oxidase activity*, which increased from 0.472 to 0.628. Conversely, the largest decrease was observed for *Cellular biosynthetic process*, which declined from 0.647 to 0.543, followed by *Small molecule metabolic process*, which decreased from 0.653 to 0.551. These results suggest that incorporating metabolomic information and the ChEBI ontology primarily reorganizes GO importance among functionally related biological processes. More broadly, terms containing the phrase *‘lipid’* increased substantially in the rankings, whereas terms containing *‘signal’* showed a modest decrease (Methods). This pattern is consistent with the greater emphasis placed by the combined model on highly informative lipid measurements^57^.

### OGM Supports Disease-Specific Transfer Learning and Downstream Analysis

Neural networks can learn representations during training on one task that are subsequently transferable to related tasks. This enables pretraining on large datasets while retaining the capacity to generalize in data-scarce settings^58^, thereby supporting downstream applications that may be difficult to achieve with linear models. This paradigm is particularly well suited to human genetics, where a small number of large biobanks provide general population data, and numerous smaller, disease-specific cohorts include rare phenotypes and additional data modalities^59^. To evaluate the transfer-learning capabilities of our model, we trained an OGM model on UKB data to predict systemic lupus erythematosus (SLE), a chronic autoimmune disease characterised by widespread inflammation. We then transferred the pretrained model to an independent single-cell dataset^60^ comprising 160 individuals with SLE and 98 healthy controls (Methods). In addition to genotype and SLE phenotype data, this cohort includes transcriptomic profiles from nine peripheral blood immune cell types, enabling us to compare genotype–transcriptome IntH values with the eQTLs identified in the original study.

After establishing that the model achieved good predictive performance in this disease-specific dataset (Figure 6a; Supplementary Table 5, Methods), we focused on identifying associations between the genotype and transcript data to better understand SLE biology. We hypothesized that the SNP and expressed gene (eSNP and eGene) that make up an expression Quantitative Trait Locus (eQTL) as described by Perez *et al.* should have stronger interactions than pairs of mismatched eSNPs and eGenes or pairs without eQTL involvement. In our analyses, we found, that IntH between genotypes and transcripts recovered established eQTL structure. Known eSNP–eGene pairs had higher absolute IntH values than control pairs, with the hierarchy known pair > eSNP-only > eGene-only > neither across all cell types (all Mann–Whitney U p < 10⁻¹⁰; Figure 6b). eSNPs were also enriched for cis interactions (Supplementary Figure 6a), and absolute IntH correlated positively with independently estimated eQTL effect sizes in all cell types except CD4 T cells (Figure 6c). IntH landscapes showed moderate similarity across cell types, consistent with a mixture of shared and cell-type-specific regulation reported by Perez et al. (Supplementary Figure 6b).

**Figure 6.**
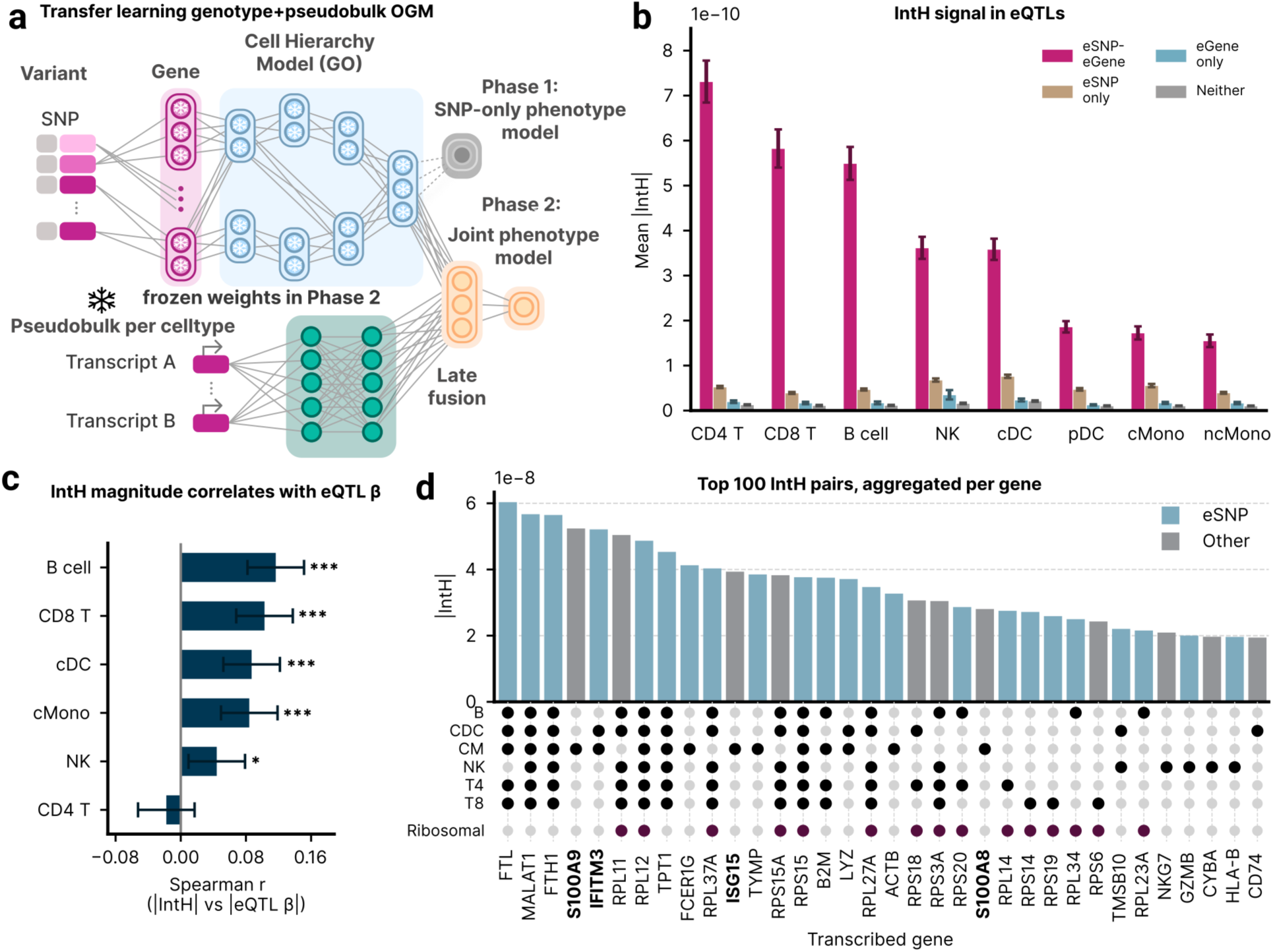
Integrated Hessians recapitulate and extend cell-type-specific eQTL interactions and recover SLE-specific immune biology. (**a**) Transfer learning and IntH analysis pipeline. The OGM encoder, pre-trained on UK Biobank data using a 100,000-variant SLE polygenic score set (PGS000754), is frozen and used to embed donor genotypes from Perez et al. (2022; 258 donors: 160 SLE, 98 healthy controls). Single-cell profiles are pseudo bulked per donor and cell type by mean-aggregating log-normalised counts. A lightweight late-fusion decoder is trained per cell type to predict SLE status. IntH scores are computed for 2,664 eSNP anchors from the Perez et al. eQTL catalog and 2,664 randomly drawn non-eSNP anchors against all 5,886 eGenes, yielding donor-averaged interaction matrices per cell type. (**b**) Distribution of mean IntH stratified by pair category across cell-type models: known eQTL pairs (eSNP anchor × eGene), eSNP-only pairs, eGene-only pairs, and null pairs. (**c**) Spearman correlation between IntH and independently estimated eQTL effect size |β|, per cell type; significant in most but not all cell types, indicating IntH magnitude does not uniformly track quantitative eQTL effect sizes across cell types. (*<0.05, **<0.01, ***<0.001) (**c**) Spearman correlation of IntH and eQTL effect sizes are positive and statistically significant for all cell types except CD4 T-cells. (**d**) Top 100 IntH pairs, aggregated per eGene, ranked by the highest IntH value per eGene. Cell types are indicated in the upset plot below, and established eSNP are color-coded. Ribosomal proteins are separately indicated in the upset plot and genes of interest not previously described as eGenes are marked in bold.

The highest-scoring variant–gene pairs included established eGenes and broadly expressed genes, but also associations not reported by Perez et al., including previously unclassified eSNPs and more cell-type-specific signals (Figure 6d). In classical monocytes, these included SLE-relevant interferon-response genes ISG15 and IFITM3^61^ and the inflammatory genes S100A8 and S100A9^62^, all supported by independent literature. Genome-wide, genes with established SLE or autoimmune associations as defined by Open Targets^63^ (see Methods) ranked significantly higher by IntH than other genes (p = 5.0 × 10⁻⁵). Together, these results show that OGM representations can be transferred to an independent cohort, recover established regulatory biology, and identify plausible disease-relevant associations beyond those previously described in its training data.

## Discussion

We propose the omnigenic model architecture for phenotype prediction from human genotypes, supporting polygenic scoring, multilabel, multimodal, and transfer-learning applications. The architecture learns multidimensional representations across increasing levels of biological abstraction defined by the GO and is inspired by the omnigenic model of complex traits proposed by Boyle et al^31^.

In its first layer, the OGM learns gene representations from individual-level genotypes, sex and genetic principal components. Both the gene layer and the early integration of covariates were critical to model performance (Figure 2e), suggesting that genotype representations benefit from being contextualized by ancestry and sex.

This deep integration of covariates represents a further step in the progression of multi-ancestry polygenic prediction described by Ruan et al.^33^, from multi-discovery models to PRS-CSx, which combines ancestry-specific summary statistics. The OGM extends this progression by using individual-level genotypes together with a continuous ancestry representation. This avoids some of the limitations of discrete ancestry assignments^35^ (Supplementary Figure 7b). In this study, discrete ancestry labels were used only to report ancestry-stratified performance, which remains essential for assessing the transferability of polygenic prediction models^64^.

Across the evaluated phenotypes and ancestry groups, OGM models matched or outperformed published polygenic scores^32^ and PRS-CSx models generated from Pan-UK Biobank summary statistics^33,34^ (Figure 2a–d). OGM-based models also generally transferred better across ancestry groups than PRS-CSx scores. Multilabel OGM models further outperformed matched single-label models (Figure 4f, g). Although additive PGS can be adapted to exploit information shared across phenotypes, such approaches remain fundamentally constrained by collapsing genetic effects into a single score^15^.

Performance nevertheless remained low in the African-ancestry group for both OGM and baseline models, likely reflecting the limited representation of this ancestry in population cohorts such as UKB^65^. Individuals of non-European ancestry comprised only 5% of the training data yet excluding them substantially reduced model performance in the more ancestrally diverse All of Us cohort (Figure 1e). This finding suggests that even modest increases in training-data diversity may improve model transferability.

Comparisons with published scores should be interpreted cautiously because phenotype definitions, age-of-onset criteria, and follow-up duration may differ even when phenotype labels appear equivalent. In addition, the large number of PGS Catalog entries available made exhaustive benchmarking impractical, and better-matched published scores may therefore exist. However, all evaluated models were tested in the same individuals using identical phecode definitions, supporting direct comparisons within our benchmark. We used phecode definitions throughout to facilitate reproducible comparisons with future studies^66^.

Related biologically structured neural-network architectures for genotype-based predictions have previously been proposed^19–21^. However, these approaches differ from the OGM in several respects, including the absence of an explicit gene layer or integrated covariates, no performance improvement over linear models, narrower task definitions, limited benchmarking against published scores, and training on substantially smaller datasets.

The advantages of the OGM architecture depend on end-to-end training using population-scale individual-level data. Although the architecture can technically accommodate more than one million variants, the associated computational cost currently makes this impractical. We therefore restricted the input to 100,000 variants, providing broad genomic coverage while maintaining practical model sizes. Our variant-selection strategy introduces an additional limitation. Variants were selected using published PGS weights^32^ and reported epistatic associations, thereby favoring established knowledge and limiting the potential to discover entirely novel disease-associated variants.

The hierarchical representations learned by OGM are also well suited to multimodal integration. Deep integration of genetic and metabolomic information substantially improved performance relative to generic neural networks trained on the same data. This is consistent with findings from methodologically distinct studies using UKB or comparable datasets^67–69^. We also observed greater synergistic benefits from combining the two modalities with the OGM architecture than with a generic neural network.

GO importance scores provide a direct form of model interpretation by identifying the biological processes that contribute most to predictive performance. Because the GO structure and gene-to-GO mappings are fixed, these analyses cannot identify entirely novel biological systems. Instead, they estimate the relative importance of predefined processes. Across our models, the highest-ranking processes showed substantial agreement with prior literature. Such interpretability may also be advantageous in eventual clinical applications, where transparent models may be preferable to less interpretable alternatives^26^. Importantly, GO importance scores are associative and do not provide causal evidence, but they can help prioritize biological systems and interactions for further investigation.

Finally, the OGM enables the *in silico* analysis of feature interactions. Interactions quantified using IntH^38^ were concordant with established epistatic relationships and known eQTL structure. The IntH analyses also identified plausible, clinically relevant interactions supported by the broader literature but not reported in the original analyses of these datasets. Across our IntH analyses, variants with high interaction scores tended to interact with many other variants rather than forming isolated high-scoring pairs. This pattern resembles the broader interaction hubs observed in systematic yeast screens, in which epistatic relationships can be investigated through targeted gene knockouts (Costanzo et al., 2016).

Together, these findings support OGM as a highly customizable and broadly applicable architecture for genotype-based phenotype prediction, as well as a strong foundation for future methodological development. We have already adapted the OGM architecture to encode transcriptomic data in NetworkVI, a framework for variational inference on multimodal single-cell data, and observed similar benefits from its multilevel representations^30^. Realizing the architecture’s full potential will likely require training across multiple large cohorts, since larger sample sizes benefit neural networks more than additive models^22^. This currently requires shared computing infrastructure, which regulatory frameworks around cohorts like UKB and All of Us complicate, but could be approached through federated learning^70^. Finally, adapting the gene layer to incorporate structural variants^71^, environmental factors^72^ or rare variants^73^ could support more comprehensive models of genotype–phenotype relationships.

## Author Contributions

J.U. and R.E. conceived the initial concept with support from N.H. and T.B.

J.U., L.A., N.H., L.H., K.N., L.E., L.K. and H.S. implemented the OGM models.

L.H. and L.K. implemented the unified datamodule udm with help from L.A. and J.U.

J.U., S.A.G, L.E., and L.A. implemented the feature selection and constructed genotype datasets.

J.U. and L.A. conducted the experiments and performed data analysis with support from L.E., L.K., S.A.G. and K.N.

L.A. implemented the Integrated Hessians analysis and adapted the OGM for application to the lupus dataset and performed the analysis with support from J.U., L.H.

J.S., S.H. and F.J.T. supported the analyses and discussion of the results.

J.U. and L.A. wrote the manuscript with support from B.W. and R.E.

All authors read, revised and approved the manuscript.

### Acknowledgements

We thank Maik Pietzner, Alice Braun, Simon Sasse, Benedict Wolf, and Tim Landgraf for helpful discussions.

We are grateful to the study participants, organizers and funders that have enabled this work:

This research has been conducted using data from UK Biobank, a major biomedical database (www.ukbiobank.ac.uk)

We gratefully acknowledge All of Us participants for their contributions, without whom this research would not have been possible.

We also thank the National Institutes of Health’s All of Us Research Program for making available the participant data examined in this study. (www.allofus.nih.gov)

Additionally, we thank the participants and coordinators of dbgap datasets phs002812.v2.p1.

We further thank the Google Research Credits Program and the Charité Scientific Computing Team and BIH project management for supporting this work.

We are grateful to Dr. Alexandra Friedrich for supporting this work.

L.A. was supported by the Helmholtz Association, as part of the joint research school Munich School for Data Science (MUDS).

## Code Availability

Code for the omnigenic model (OGM) is available on GitHub under MIT License https://github.com/juzb/ogm

Code for the unified datamodule (udm) is available on GitHub under MIT License https://github.com/luisherrmann/udm

## Data availability

Individual level records are available through the UK-Biobank and All of Us. Aggregate data is available in the Supplementary files and on GitHub alongside the code. The genotypes of the SLE cohort from Perez et al. (2022)^60^ are available at dbGap accession number phs002812.v1.p1, the scRNA-seq data is available at https://cellxgene.cziscience.com/collections/436154da-bcf1-4130-9c8b-120ff9a888f2.

## Ethics oversight

Ethical approval for this study was obtained from the Ethics Committee of Charité Universitätsmedizin Berlin.

The UK Biobank has obtained approval from the North West Multi-centre Research Ethics Committee.

The All of Us program has obtained approval from the All of Us Institutional Review Board.

## Supplementary files

We provide larger tables and figures in a supplementary file. This includes

1. Full model performance data
2. Full GO importance values for each figure
3. Complete GO importance values across the entire model for each figure.
4. Variant lists used to create our genotype datasets
5. Mapping of NMR metabolites to the ChEBI ontology

## Methods

### Phenotype- and Covariate Processing

The UK Biobank population is 46 % female and has a median age of 76.

We use genetic sex (UKB field 22001) and genetic principal components (UKB field 22009) as covariates in our models and PGS baselines. Sex is kept as a binary variable, and the genetic PCs are used as provided by the UKB^65^ in the UKB cohort. In the AoU cohort, PCs based on UKB eigenvalues (UKB resource 149744) are standardized to be in the range of values as in the UKB.

The All of Us population is 62 % female and has a median age of 60.

In the AoU cohort, we only include individuals in which the sex at birth entry and dragen-predicted sex ploidy match^74^, which excludes < 2% of the population.

### UKB Genotypes

We work with the imputed genotypes in the UK-Biobank. These are available for practically the entire population and contain 93,095,624 variants in the .bgen-format in genome version GRCh37. The genotypes are stored as soft dosages.

### AoU Genotypes

We work with the short-read whole genome sequencing data from AoU release v7, which contains genotypes for 245,394 individuals with matching sex and available phenotypes. Specifically, we use the ACAF-threshold-subset, which contains only variants with an allele count > 100, or MAF > 0.01 for any ancestry group. While this contains fewer variants, it is also available in .bgen format (which the entire WGS data is not), massively simplifying our workflows. The AoU genotypes are in genome version GRCh38.

### Harmonisation of genotypes

Since we require consistent variant sets across the UKB and AoU cohorts, we crossmapped^75^ all UKB-variants from GRCh37 to GRCh38. We then determined the intersection of the variant sets and retained 18,880,820 variants that were usable for this work. While genotyping arrays exist in the AoU cohort, no imputed genotypes are available directly. Thus, working with imputed array genotypes on the UKB and WGS genotypes in AoU presents a compromise, however to the disadvantage of our models, posing a more challenging external validation.

### Genotype datasets

Genotype datasets for training large neural networks must support fast sample-level access. This is orthogonal to the requirements for GWAS which require fast variant-level access and therefore work well with formats like the .bgen format which compresses genotypes along that axis. To allow for simple memory-mapping dataloaders, we write the genotypes to a numpy matrix that contains the required variants for the entire population. These genotype matrices are annotated with tables containing the individual IDs (EID in the UKB, research_id in AoU) for the sample axis, and Ensembl Gene IDs (among other annotations) on the variant axis.

We have multiple, partly conflicting requirements for the selection of variants to be included in our dataset. Firstly, the variants should be informative of phenotypes. Secondly, they should not be highly correlated as this would decrease the effective dimensionality of the data. Notably, the model can deal well with correlated inputs, but since we are limited on the input dimensionality for practical reasons, it is desirable to have a high information content within them. Thirdly, the variant should cover the entire genome, to enable the prediction of multiple diseases based on the same dataset. We achieve these goals by picking the top variants from published PGS, excluding those using UKB or AOU data to avoid re-using the same data for feature selection, model training and model evaluation. To also cover the entire genome, we ensure that all genes are represented by roughly the same number of variants (Figure 1d).

To further investigate the effect of epistasis, we replace the lowest ranked variants with variants previously reported to show epistatic effects (see “Feature attribution and epistasis”). These datasets are used for all models of cardiovascular phenotypes. A subset of these epistasis variants is not directly available in the All of Us genotypes. For the integrated Hessians and integrated gradients experiments which we run on the UKB data, we therefore disable this filter.

For the SLE experiments we used the same variant selection pipeline, but used only the weights of PGS000754, a PGS for SLE^76^, skipping the initial multi-PGS steps.

### PGS calculation

We used the plink2 score function to calculate polygenic risk scores on the UK-Biobank imputed genotypes and generally observe > 90 % of variants covered for published polygenic risk scores. Similarly, on the AoU cohort we use the .bgen files of the ACAF genotypes, which get > 90 % of variant coverage as well. When using scores from the PGS-catalog^32^, we rely on the harmonized versions of the PGS provided, that contain coordinates in GRCh38 as well as GRCh37. Since genotypes are available in per-chromosome .bgen files we sum the scores across chromosomes and normalize by the count of available alleles for each individual.

### PGS baselines on UKB data with PRS-CSx

Custom PGS baselines were computed using the PRS-CSx framework^33^ and UK-Biobank summary statistics from the Pan UKBB project^34^. Specifically, we fix parameters a=1 and b=1/2, (the Strawderman-Berger Prior), and for IHD, T2D and schizoprenia we run a sweep across the global shrinkage parameter phi from 1e-1 to 1e-10, which controls the polygenicity of the model. For the baselines across cardiovascular phenotypes, we require substantially more models, and sample the phi parameter coarser, specifically we test phi=1e-2, 1e-4, and 1e-6, covering the recommended range. To ensure that the PGS are AoU compatible, we restrict them to the intersection of UKB and AoU variants when running PRS-CSx. We pick the best model on the validation set defined for our models. For the IHD, T2D and schizophrenia baselines we manually selected the PanUKBB entries phecode-411, phecodes-250.2, and phecode-295.1. For the multilabel baselines, we automatically mapped the phecodes entries to phecodeX labels we used for model training.

### PGS baselines with covariates

To compare the PGS fairly to models that include covariates like sex and genetic PCs, we take the pre-computed individual scores, concatenate the pre-processed covariates, and fit a logistic regression for the respective phenotype using the scikit-learn package^77^. The covariates are genetic sex and the first three genetic principal components, as well as optionally age. We then save the fitted logistic regressions and normalisation factors to files and upload them to the AoU workspace to run there. Thus, when we report an evaluation for a PGS with covariates on the AoU cohort, the entire setup (PGS, logistic regression) is only informed by UKB data and applied to the AoU data as is.

### Genetic principal components

Since both our models and our PGS baselines with covariates utilize genetic principal components, we require a common principal-component space between UKB and AoU. We chose to work in the UKB PC space, so that only the UKB informed our setup, and we could theoretically transfer the entire setup to a completely different cohort instead of AoU. Specifically, we use the PCs provided in UKB field 22009, and download the eigenvalue files provided. We upload these to the AoU population after mapping them to GRCh38 and use the plink2 score function to compute the AoU genetic PCs in the UKB PC space. While the two PC spaces look different, we see high correlations, especially in the first few PCs, and a clear separation of ancestries (Supplementary Figure 1), confirming that the PCs computed this way can be used to approximate ancestry consistently in the two cohorts. This approach is easily applicable to any new cohort, more specifically an existing model trained on the UKB can be transferred to a new cohort just using the UKB eigenvalues to construct the appropriate PC space for the model.

### Definition of subpopulations

#### UK-Biobank

Since our models are being trained and hyperparameters selected on UKB data, we have multiple requirements for the definition of subpopulations. Firstly, we require a training dataset that is informative for the entire population (train). Then we require a dataset to evaluate models on during training, and to select hyperparameters for the final model (valid). Finally, we need a testing population on which we can determine the final model’s performance on the UKB population (test). We extend this with a second testing population of non-white British ancestry (non-white test).

To achieve this, we first exclude individuals without sufficient genotyping quality, or missing EHRs (Figure 1c). Then we take the white British subpopulation (Field 22006), excluding anyone with kinship up to 3rd degree based on the kinship file provided by UKB (Field 22021), and randomly assign 60 % to the train, and 20 % to valid and test splits each without overlaps. The training data is then extended by the individuals with kinship, this introduces more population structure (about which our models do not make assumptions), but also increases the size of the train dataset substantially. In a last step, the non white-British population is randomly split in two sets of equal size, one added to the train dataset, and the other one is then the non-white test population. This setup ensures that there is no leakage between the train population and the other populations through kinship. This would occur, e.g. if one sibling were in the train population, and another in the test population.

Our training dataset contains 338,499 individuals. The EUR training only dataset (Figure 2d) contains 294,344 individuals, and the model selection dataset contains 54,296 individuals. The EUR test dataset contains 54,296 individuals, and the non-EUR test dataset contains 28,779 individuals.

#### All of Us

For the AoU cohort, we are only interested in evaluating our models, thus the number of samples per subpopulation becomes the focus, as it ultimately determines the size of the confidence intervals for our metrics. We use the AoU-provided ancestry assignments (which are based on genetic PCs) to distinguish between subpopulations of different ancestry. This also makes our results more easily comparable to other works using the AoU-provided ancestry definitions. Additionally, we use the same sex definition as in the covariates described above.

The external test datasets contain 102,633 EUR ancestry, 30,788 AMR ancestry, 36,116 AFR ancestry, 3,253 EAS ancestry, 2,069 SAS ancestry, and 676 MID ancestry participants.

#### Unified datamodule

To handle genotype, phenotype, and covariate data from different files, for different sample sets, and possibly subset to specific columns, we rely on a custom udm python module that we publish alongside the OGM code. It reads data from different formats, including .csv files, .feather files, .npy files and processes them into pytorch dataloaders that we use to train our models.

#### Model implementation

The models are implemented in pytorch and can run on a single V100 GPU, and trained within 24h on the full UKB training data. They typically have 5M-10M trainable parameters and are trained like a multilayer perceptron for the binary prediction task of disease risk prediction. We use the binary cross entropy as the loss function and utilise early stopping to avoid overfitting. Inference in AoU for 250,000 individuals runs in 20min on a V100 GPU, or within up to 2h on CPU, which is crucial for application in trusted research environments without GPUs.

Variants are assigned to genes using annotations from SNPEffv2, and genes are mapped to the gene ontology (version from 2024-08-05) via the mapping provided from the gene ontology (version from 2024-08-05).

Genes that could not be mapped to the GO in this way are mapped to a special ‘GO-Unknown’ module in the network. It is a configurable multilayer perceptron that typically has 2 layers with 1000 hidden units in the intermediate level and 20 hidden units on the final level. It directly connects to the root node of the GO.

Within the GO we work with the biological process namespace unless otherwise stated and use the ‘is_a’ relations. We provide comparisons with the cellular component and molecular function ontologies in Figure 2e, confirming the biological process ontology to lead to higher performance. It also provides a more intuitive hierarchy for providing increasingly abstract genotype representations, and linking them do disease-biology.

We provide a simple gene model that just maps variants to genes as described above and then connects them to the GO. Additionally, we developed a covariate-input gene model that concatenates a subset of covariates (here genetic sex, and genetic PCs 1-3) to the input for each gene. Thus, the gene representation is based on these covariates and the variants associated with that gene. We use this version with covariate inputs unless otherwise stated.

##### Blocksparse layers

While it is theoretically possible to represent each GO term with a torch.nn.linear object and pass their inputs and outputs according to the connectivity of the GO, this is not feasible in practice. It would end up with thousands of individual objects that each only perform small calculations, typically having less than 1000 parameters. Instead, each transition from one GO-Level to another can equivalently be expressed in a single torch.nn.linear with a high degree of sparsity, that follows a block-sparse pattern. This requires much fewer objects and hence reduces overhead but costs some additional overhead for the sparse matrix multiplication instead of dense matrix multiplications. Since the GO contains connections that jump over levels (e.g. from level 5 directly to level 3), this representation requires L x (L-1) / 2 layers for a GO with L levels. Specifically, it contains layers for the transitions from L2 to L1, L3 to L1, L3 to L2 and so forth. The resulting representations of transitions arriving at the same level can then just be added up, e.g. the one from L2 to L1 and the one from L3 to L1. Since this implementation leads to irregular sparsity patterns with high variance in the number of non-zero elements in each row and columns, we apply the commonly used Kaiming uniform initialisation^78^ for each block individually, which speeds up model training. We optionally extend the Blocksparse Layer by a residual option that is intended to allow the layer to compute a linear, and a non-linear transformation of its input and sum these up. This is inspired by the resnet architecture, which is not directly applicable in the case of our models because it would require input and output dimensions of the layers to be identical. Including this residual connection is treated as a binary hyperparameter.

##### Prediction Heads

To ensure modularity and flexibility, take the genotype (and optionally covariate) embeddings and pass them to one or more prediction heads. These are small neural network modules that can take additional inputs (like further covariates), and all predict the phenotype. This allows for the simultaneous optimisation of multiple sub-models and allows for the optimisation of the main model via different heads. We use only one head for phenotype prediction throughout the manuscript with the exception of the late covariate fusion ablation (Figure 2e) where we have a genotypes+covariates head; and the exception of the metabolomics models where we provide the metabolites both through the GO-CheBI network, and akin to a residual connection, directly to the prediction head.

#### Training

Models are optimised against a binary cross entropy loss using the adamw^79^ optimiser with weight decay 0.1 and are trained for up to 30 epochs using early stopping based on the valid sets AUROC to address overfitting. We use a batch size of 64 or 128 if GPU memory permits, batch normalisation. Training takes around 24h on a V100 GPU.

#### Hyperparameter Optimisation

Our models come with the standard hyperparameters of neural networks and introduce a few new ones specific to the architecture. The GO depth controls how many levels of the GO are represented in the model, and because the lower levels have exponentially more terms, heavily influences the number of trainable parameters. The GO size and gene size parameters control the number of neurons each GO, or gene terms is represented by. Additionally, we allow for the final layer of the GO to have a larger dimensionality, controlled by the latent size parameter.

Genes not mapped to GO terms are mapped to a single GO-Unknown module that is a small standard neural network with controllable number of layers and neurons per layer.

Optionally, we can normalise the model inputs, which is controlled by a binary flag, and implemented as z-scoring the genotypes before passing them to the first layer. Similarly, to regularise the model, we can introduce controlled gaussian noise to the genotypes which is controlled through a noise-parameter. Hyperparameters are optimised using the ax framework^80^, specifically using the Gaussian Process + Expected Improvement Generation strategy, set to maximise performance in the valid split.

#### Interpretability

The domain knowledge-driven architecture allows for the inference of a metric representing the importance of a group of nodes, such as a GO term, for the model performance. Logistic regressions are fitted using the activations in relation to the sample labels, as previously illustrated by Ma *et al*^28^. The significance of each GO term is quantified by the area under the receiver operating characteristic curve (AUROC) from the logistic regression models and referred to as GO importance.

For multilabel experiments, including the metabolomics experiments, the higher memory burden required subsetting the samples used to 50 % of the population, which we did in an unbiased manner to allow for GO importance analysis for all phenotypes.

Our models typically contain thousands of GO terms, which necessitates selecting subsets for the figures here. To provide a reproducible selection process, we implemented two algorithms to produce candidate subsets, which we show in full in the supplementary files, and manually subset them to reduce redundancy if necessary (Figures 3c, 5c, Supplementary Figure 2, 3). For example, in long linear chains without branches, we only show the most specific term, or we only show either synthesis or catabolism, either transport or localisation, while trying to maintain the coherence of the hierarchy. We provide full go importance values for the models in the supplementary files.

The first algorithm (top path) starts at the root node of the ontology and greedily descends into the term with the highest GO importance at each level. Then, all parent nodes of all selected terms are included as well, to provide the full context. For terms with only one parent, this results in a linear graph, so we include a parameter to control the number of the highest-ranking terms included per layer, this is set to 2. For the model with metabolomics-integration (Figure 5d), we use only the highest ranking terms on this path, and do not include all parent-terms of the terms involved. Otherwise, the high connectivity of low-level ChEBI-terms into the GO produces unreadable graphs.

The second algorithm (top leaves) picks the highest-ranking leaf nodes and includes all their parent terms up to the root node. We again disable this parent inclusion for the mtabolomics-integration models.

#### Feature attributions and Epistasis

To evaluate feature importance in our model, we calculated Integrated Gradients^38^ (IG) for all SNPs in the dataset. To investigate variant-variant interactions, we additionally curated 39 SNPs corresponding to 14 literature-reported epistatic pairs associated with coronary heart disease (CHD, n=5 pairs), blood pressure (BP, n=2 pairs) and body-mass index (BMI, n=7 pairs), identified by searching for evidence of epistasis in connection with the terms “coronary heart disease”, “diabetes type 2”, “BMI”, and “blood pressure”^81–88^. These pairs represent an excerpt of the broader collection of epistatic interactions reported in the literature. We provide the specific pairs in Supplementary Table 1 alongside their IntH values.

To compare the literature-derived SNPs against a background, we additionally selected a non-overlapping sets of 39 background SNPs which was randomly selected (seed=42). IntH were computed jointly for all 39 literature SNPs and the respective 100 background SNPs as queries against all 100,000 modeled SNPs, separately for each background option. We use the mean of absolute individual IntH values as the IntH matrix for each option. Variant importance, for IG was similarly calculated as the mean across all samples absolute values.

We tested for non-additive signal in two complementary ways. First, for the full 39-variant set, we extracted the 39×39 Epi×Epi sub-matrix (741 upper-triangle pairs), the 39×39 Epi×Random matrix, and the 39×39 Random×Random sub-matrix. We compared Epi×Epi against Epi×Random using a one-sided Mann–Whitney U test; Epi×Epi against Random×Random likewise. We adjusted the p-values for the three tests, using a Bonferroni correction. Second, for the 3 CHD epistasis pairs with at least one GO term annotated specifically, we compute the percentile of their IntH in the full interaction set, including other Epi×Epi, Epi×Random and Random×Random. IntH computations used num_samples=10 integration steps and were averaged over 1024 individuals.

#### Comparison of Integrated Gradients with a published polygenic risk score

To benchmark variant-level model attributions against an orthogonal, literature-established signal, we compared IG against the variant weights of PGS000013^10^. We found a matching PGS weight for 72,461 of the 100,000 modeled SNPs (72.5%). SNP-level concordance between mean |IG| and |PGS weight| was assessed by top-k set overlap at logarithmicicall spaced thresholds using one-sided Fisher’s Odds ratio, comparing observed Jaccard indices against their chance-expected values.

Candidate loci for detailed visualization (Supplementary Figure 4) were selected by aggregating SNP-level |IG| and |PGS weight| per gene (sum across each gene’s matched SNPs, as above), independently ranking genes by IG-sum and PGS-sum percentile, and taking the minimum of the two percentiles per gene. Genes were sorted by this joint percentile and the top-ranking entries were manually reviewed; six widely-recognized ischemic artery disease and lipid-trait loci (APOE, CDKN2B-AS1, APOE, CELSR2/SORT1, PHACTR1, LDLR) were selected for visualization in Supplementary Figure 3.

#### Ablation Studies

To systematically investigate the importance of the gene layer, the GO graph, and the overall structural design of the model for its predictive performance, we conducted a series of ablation studies for the IHD phenotype. For ablation models changing the architecture, comprehensive hyperparameter optimization was performed to ensure fair comparison with the full model. The specific hyperparameter settings for each model are provided in the supplementary materials (Supplementary Table 2). For direct comparisons with fixed hyperparameters, we only modified the specific ablation condition. This includes ontology namespace (to cellular component and to molecular function), the training samples (to EUR only, and to EUR only excluding kinship), and the randomised variant-to gene mapping. The ablation experiments confirm that the OGM architecture outperforms standard neural networks, benefits from the gene model and the GO-model and uses the biological information of the GO rather than its sparsity, by shuffling the variant-to-gene mapping. Additionally, we confirm that the GO biological process ontology is better suited for this application than cellular component or molecular function. Finally using only European ancestry training data or additionally excluding individuals with kinship from the training data substantially decreases performance. In separate comparisons including covariates we confirm that their integration into the gene model is beneficial, and that the inclusion of the epistatic variants (Methods: Feature attribution and epistasis) can improve model performance measurably.

#### Multilabel experiments

Multilabel phenotype datasets were constructed by including all 3-digit phecodes (up to the “.”) from the cardiovascular chapter (CV) with more than 1000 cases within the UKB dataset without any further filtering. Phecode CV 402 (Elevated blood pressure reading without diagnosis of hypertension) does not appear in AOU and is therefore not evaluated, all others do at least 300 times. The most common phecodes in AoU is CV 401 (Hypertension). Loss and metrics functions are replaced by macro averages across all phecodes for training purposes, and per-class performances are reported like they are for single-phentoype models.

PGS from the PGSCatalog use different phenotype definitions that are not trivial to map to phecodes. We used the best matches available based on the provided phenotype descriptions. In some cases, a direct match could not be found. To still provide context of how well the respective phenotypes might be predicted based on genetics, we looked for related phenotypes. For most phenotypes, multiple PGS existed, there we picked the one with best reported performance in European ancestry populations, in line with our model selection strategy for the OGM models and PRS-CSx baselines. This is complicated by inconsistent reporting across different metrics, phenotype definitions and evaluation cohorts. In some cases, we evaluated multiple candidate PGS in the UKB population, and picked the best one for evaluation in the AoU cohort. Due to the large number of PGS for cardiovascular phenotypes, it was impossible to do this exhaustively. Reported performance should thus be read as the performance of a comparably well-performing PGS, but not necessarily the best one overall. Where possible, we evaluated multiple PGS in the UKB population and selected the highest performing one. A detailed mapping of phecode to PGS ID and the corresponding UKB performance values are given in Supplementary Table 4.

To compare models across phenotypes we performed one-sided Wilcoxon rank-sum tests in each ancestry, for the three baselines. p-values were adjusted using the Holm-Bonferroni method, treating the tests for each ancestry as a family of tests.

#### Metabolomics

In an extension of the original model, we incorporated metabolomics data as an additional input alongside genetic data. Circulating metabolite measures were obtained from the UK Biobank Nightingale Health NMR platform, comprising 249 biomarkers per sample (absolute concentrations of lipids, lipoprotein subclass particle concentrations and compositions, fatty acids, amino acids, ketone bodies, glycolysis-related metabolites and inflammation markers, together with derived ratios and percentage measures). Only samples from the baseline visit were retained. Each biomarker was standardized within spectrometer: samples were grouped by their measuring spectrometer, and for every biomarker we subtracted the spectrometer-specific mean and divided by the spectrometer-specific standard deviation, yielding per-instrument z-scores. Where a participant contributed more than one qualifying sample, the standardized values were then averaged per participant, producing one row per individual. The metabolomics values were mapped manually by the authors to the **ChEBI Ontology** (release of 27 May 2024, see Supplementary Table ?), and the ChEBI Ontology was further linked to the Gene Ontology using the cross-references provided within the Gene Ontology database^27^ (go-plus.owl, release of 24 April 2024). Metabolite levels change substantially over time, and dependent on diseases. We thus restrict our predictions to a 10-year window after the first assessment centre visit where the blood draw for the metabolomics took place. Phecode occurrences after the 10-year cutoff are ignored entirely. In the evaluation setting, we further exclude individuals that have the specific phecode documented before the first assessment centre from the metric computation.

To assess whether the predictive benefit of combining genotypes and metabolomics depends on the ChEBI-informed fusion specifically, or simply on making both modalities available to the model, we trained a generic neural network with genotypes and metabolomics data available in the same setting.

To quantify the change in GO importance attributable to metabolomic fusion for the heart failure model (Figure 5c), we compared its GO-importance values against those from the genotype-only cardiovascular multilabel model (Figure 4), restricted to the GO-term universe shared between the two models, analogous to the matched-universe approach used in the GO-disease association analysis above. As this genotype-only comparator was trained on the general (not 10-year-incident-restricted) prediction task rather than the exact metabolomics-matched setting, this comparison is indicative rather than a fully controlled ablation. To compare GO importances between the genotype-only model and the model including the metabolomic data, we first sorted the GO-importance values separately and assigned a rank. We computed a relative rank ranging from 0 to 1, by normalising this rank with the number of GO terms, to adjust for different term counts in the two models. After merging the two tables, we computed the difference in this relative rank and provide the highest and lowest values, as well as mean values for subsets of terms including specific phrases.

#### GO-Disease Association

To evaluate the specificity of our GO importance values, we performed a quantitative analysis of the associations between highly important GO terms in our model and disease biology. We benchmarked GO importance against literature-curated disease-GO-term annotations from two databases: the HPO2GO dataset by Dogan et al.^89^ and the CTD (Comparative Toxicogenomics Database) phenotype–biological-process associations^90^. A disease’s annotated GO-term set is the union of terms reachable from either database, intersected with nine HPO- and MESH-derived disease-vocabulary mapping schemes plus their union (“COMBINED”), restricted to GO biological-process terms.

As an independent baseline, we computed an over-representation analysis (ORA) of the GO biological-process ontology using WEBGestaltR^91^ (Fisher’s exact test, FDR-BH and Bonferroni-corrected), testing the gene set implicated by each disease’s published PGS (genes harbouring a PGS variant, weighted by |effect size|) against the GO-annotated genome as background. We report over-representation analysis (ORA) results based on FDR-BH-corrected significance on this PGS-selected gene set as our baseline for IHD and T2D.

For each candidate score (OGM importance or an enrichment baseline), we computed the enrichment of disease-associated terms in the top x % of the candidate scores from 0.3 % up to 10 %. We provide 95 % CIs by bootstrapping the enrichment calculation 200 times and using the 2.5 and 97.5 percentile values as the CI.

#### Transfer learning and fine-tuning on independent SLE cohort

To evaluate whether the OGM generalises to an independent single-cell dataset and to enable interpretability of genotype–transcriptome interactions in a disease-relevant context, we fine-tuned the OGM latent representation on data from Perez et al.^60^, a single-cell RNA-sequencing study of 258 donors (160 systemic lupus erythematosus [SLE], 98 healthy controls) profiled across nine peripheral blood immune cell types. Of the 261 donors in the original Perez et al.^60^ dataset, 3 were excluded during genotype quality control (upstream of SNP extraction), leaving 258 donors.

The OGM was trained on a 100,000-variant SLE polygenic score set (PGS000754^76^) selected as described previously and intersected with the genotypes for the Perez et al. cohort using chromosome:position coordinates. Gene expression profiles were pseudobulked per donor and cell type by mean-aggregation of log-normalised counts. The genotypes and single-cell data was loaded, preprocessed via the DonorData structure and fed to the late fusion model via the MILDataset in cellink^92^.

Donors were split by sample-processing batch: three of four processing batches (batch1, batch2, batch3) were used for training and the fourth (batch4) was held out entirely for evaluation, for all downstream models. This was necessary because a random donor-level split left SLE status confounded with processing batch in this cohort (donor-level disease prevalence varied sharply by batch), which inflated classification performance independently of genotype or transcriptome signal; the batch-based split removes this confound. Held-out test donors (n=83) were drawn from a single batch never seen during training; the remaining 175 donors (3 batches) were used for training.

The OGM encoder (all parameters) was frozen throughout fine-tuning; the prediction decoder head was removed. In place of a single fused prediction head, we used a dual-head additive fusion decoder: a main head received the concatenation of the frozen OGM genetic latent and an embedding of the pseudobulked single-cell profile (learned by a two-layer neural network), and a second, genetics-only auxiliary head was trained in parallel on the genetic latent alone, with its logit added to the main head’s logit (scaled by a learnable weight). Total training loss was the main head’s binary cross-entropy plus a weighted auxiliary loss term on the genetics-only head’s prediction. This architecture was adopted to ensure both modalities could be used effectively for SLE prediction. Separate models were trained for each available cell type (B cell, CD4⁺ T cell, CD8⁺ T cell, classical monocyte, non-classical monocyte, cDC, pDC, and NK. Training used AdamW (lr = 1×10⁻³, weight decay = 1×10⁻⁴) with binary cross-entropy loss and early stopping (patience 5 epochs). Reliance on genetic input was assessed for every trained model using a zeroing the genetic latent ablation on the held-out batch, which reduced test AUROC relative to the full model in the majority of cell types (Supplementary Table 5).

#### Integrated Hessians for SNP×gene interaction scoring

Integrated Hessians^38^ estimate the pairwise interaction between input features in a neural network as the integral of the second-order mixed partial derivative of the output with respect to two features along a straight-line path from a baseline (zero) to the observed input. For our model *f*, the IntH score for SNP *i* and gene expression feature j in donor *d* is:

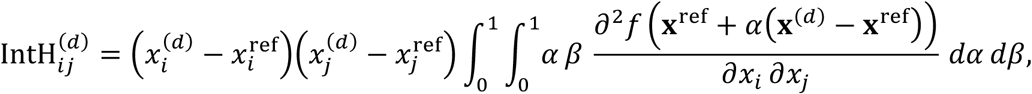

approximated by a Riemann sum over *m* = 10 steps.

IntH scores are computed for each of *n*_variant_ SNP variants against all *n*_gene_ pseudobulked gene expression features per donor, yielding a tensor of shape (*n*_variant_ × *n*_donor_ × *n*_gene_) per cell-type model. SNP variants comprise (i) all eSNPs from PGS000754, and (ii) an equal number of randomly drawn SNPs from the remainder of the array. Population-level IntH statistics are computed as a weighted mean across donors, with per-donor weights proportional to cell count, and stratified by disease label (SLE / control).

Integrated Hessians computation takes up to 48h on H200 GPUs for 140 query variants and 1280 individuals.

#### Integrated Hessian scoring and enrichment analysis

For each cell type, population-mean absolute IntH matrices were computed from the fine-tuned OGM by averaging Integrated Hessians over donors. To validate IntH scores as a proxy for regulatory activity, three tests were applied. (1) Spearman correlation between gene-level mean absolute IntH and absolute eQTL effect sizes β was computed per cell type. (2) Mean IntH was compared between eSNP and non-eSNP variants, and between eGene and non-eGene targets, using one-sided Mann–Whitney U tests. (3) Pairs were stratified into four classes (known eQTL, eSNP-only, eGene-only, null) and mean IntH was compared across strata.

For the cis/trans analysis a 1 Mb window centred on each SNP was used to classify target genes as cis or trans; the per-anchor cis/trans ratio was computed as the mean IntH at cis genes divided by the mean IntH at trans genes. Novel candidate pairs were defined as the highest-IntH anchor–gene pairs excluding catalogued eQTL pairs and a small set of anchors with disproportionately high marginal IntH across all genes (top 0.5% by mean IntH, excluded to prevent a small number of anchors dominating every cell type’s candidate list). A gene-capped ranking, restricting each gene to its single highest-IntH novel pair, was used to avoid recurrent genes crowding the candidate list (FIgure 6c). SLE-differential IntH was computed as IntH(SLE) − IntH(ctrl) for each pair, averaging IntH separately within SLE and control donors; statistical comparisons used two-sided Mann–Whitney U tests.

#### OpenTargets disease-association analysis

To assess whether IntH-prioritised genes are enriched for disease relevance, candidate genes were queried against the OpenTargets Platform^63^ GraphQL API for their top five associated diseases and association scores, and against the GWAS Catalog^93^ REST API (candidate SNPs) for trait associations. A gene or SNP was flagged as SLE/autoimmune-associated if any of its top five OpenTargets disease names matched “lupus” or “autoimmune”.

A genome-wide IntH-based importance score was computed per gene as the maximum IntH across all anchors and all ten cell-type models, after excluding catalogued eQTL pairs and high-noise anchors (as above). Genes with a confirmed SLE/autoimmune OpenTargets association (n=264) were compared against all other genes (n=5,622) by one-sided Mann–Whitney U on this IntH score.

## Supplementary Figures

**Supplementary Figure 1.**
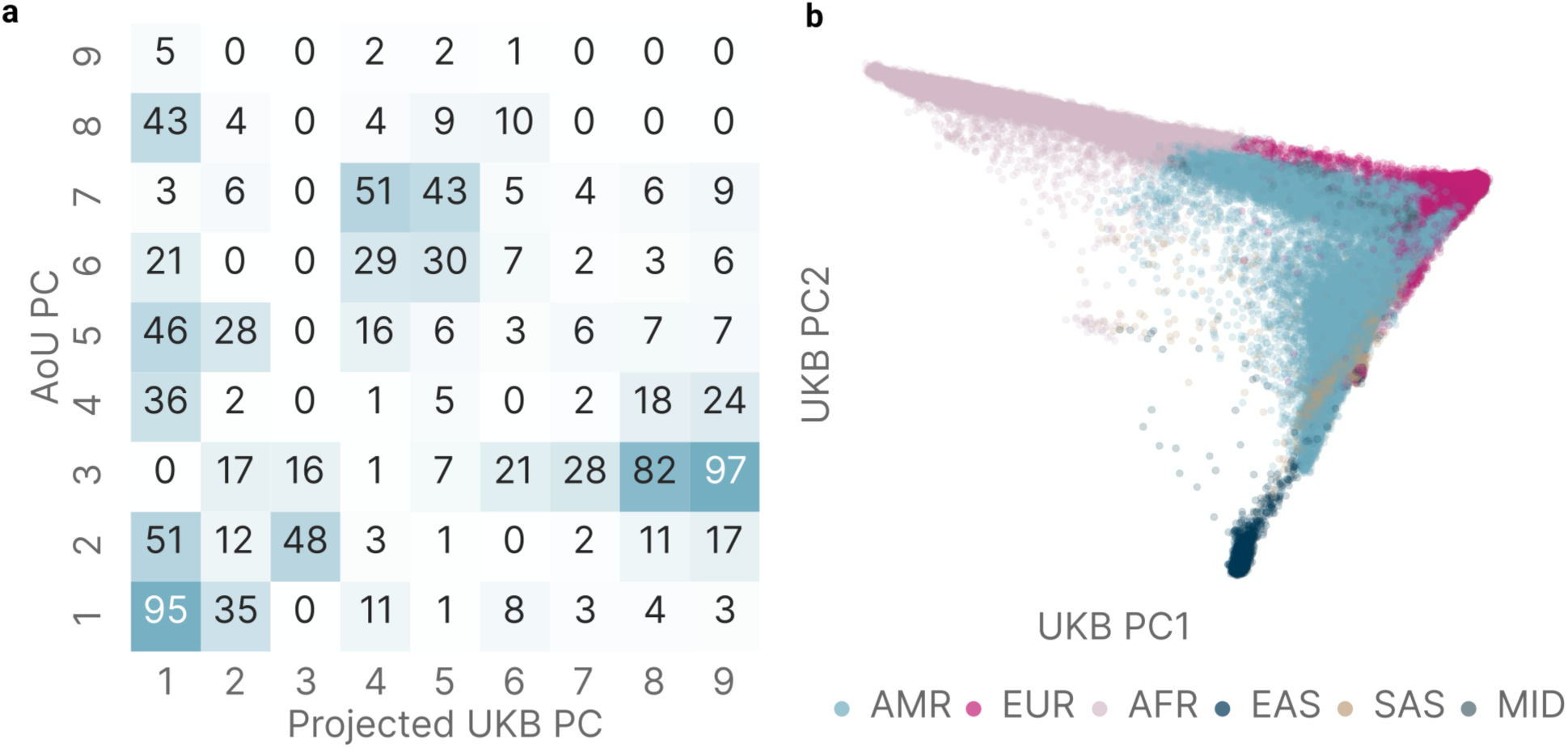
Projection of AoU samples into UKB PC space. (**a**) Absolute correlations (r2) between UKB-projected and native AoU PCs show strong overlap between the two PC spaces. (**b**) Visualisation of the first two PCs based on UKB eigenvectors in AoU. Subpopulations are clearly resolved.

**Supplementary Figure 2.**
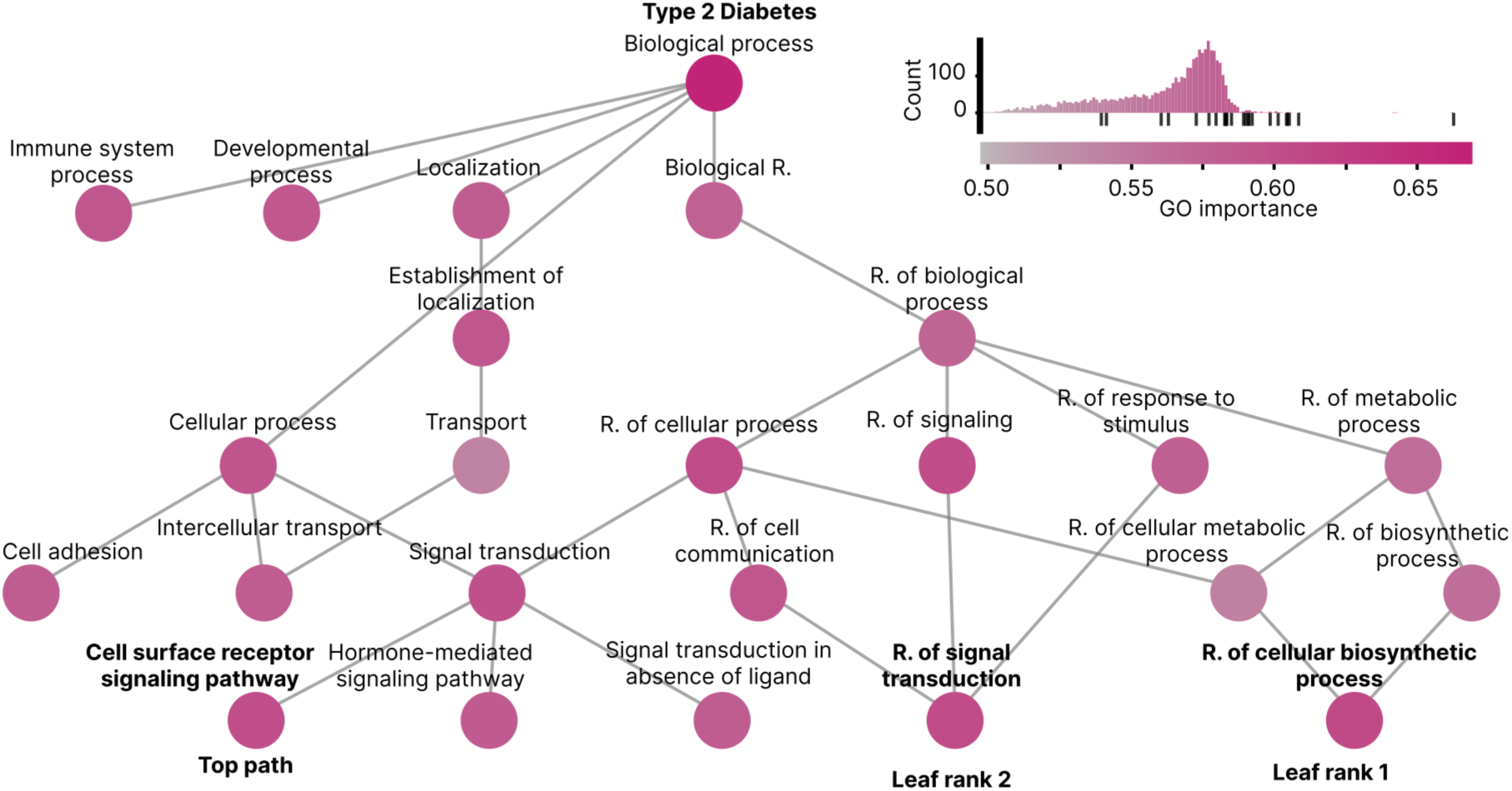
Interpretability for type 2 diabetes. GO importances for a single-label T2D models are shown for the top path, and top 3 leaf nodes, including all their parent terms with no manual exclusions.

**Supplementary Table 1.**
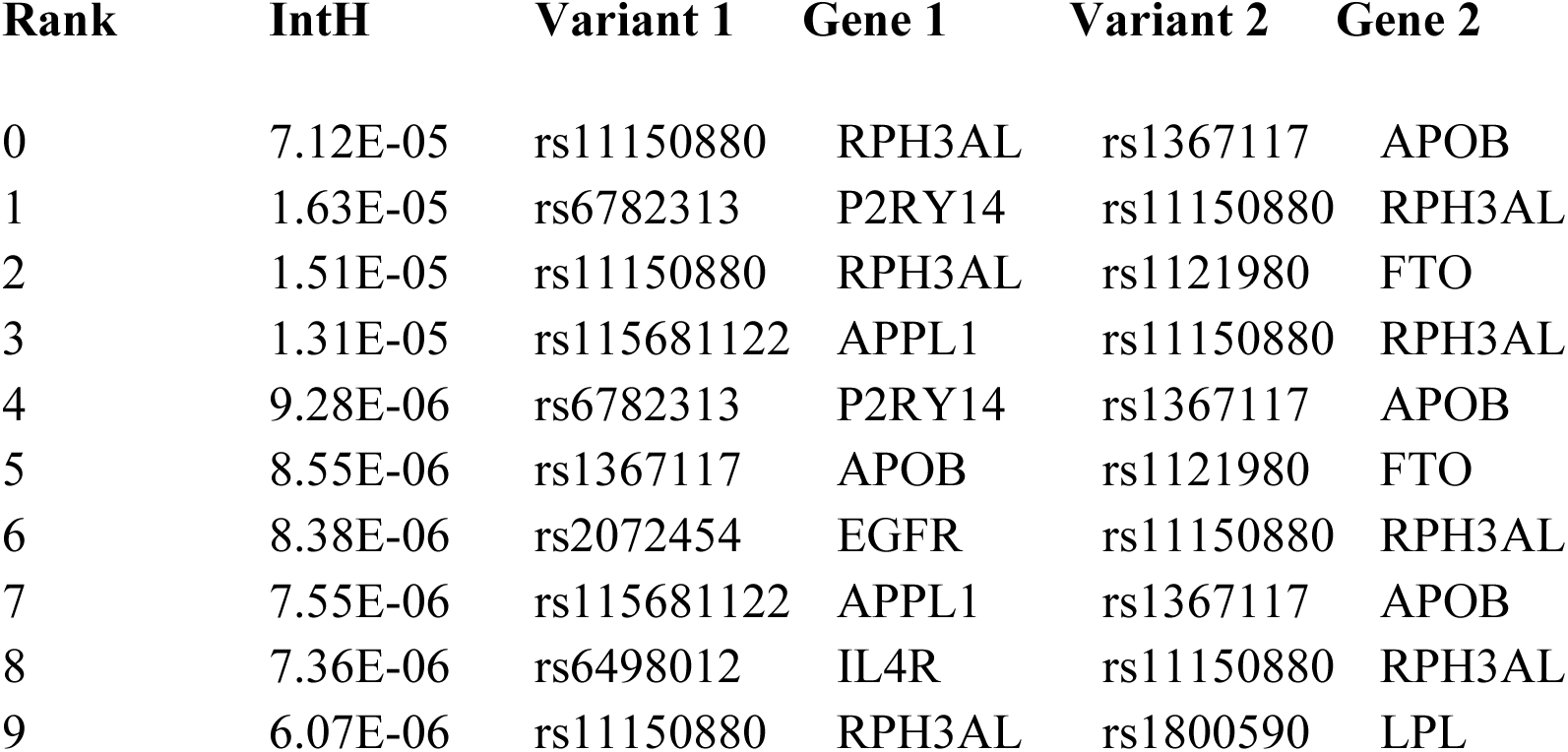
Top variant-pairs by Integrated Hessians.

**Supplementary Figure 3.**
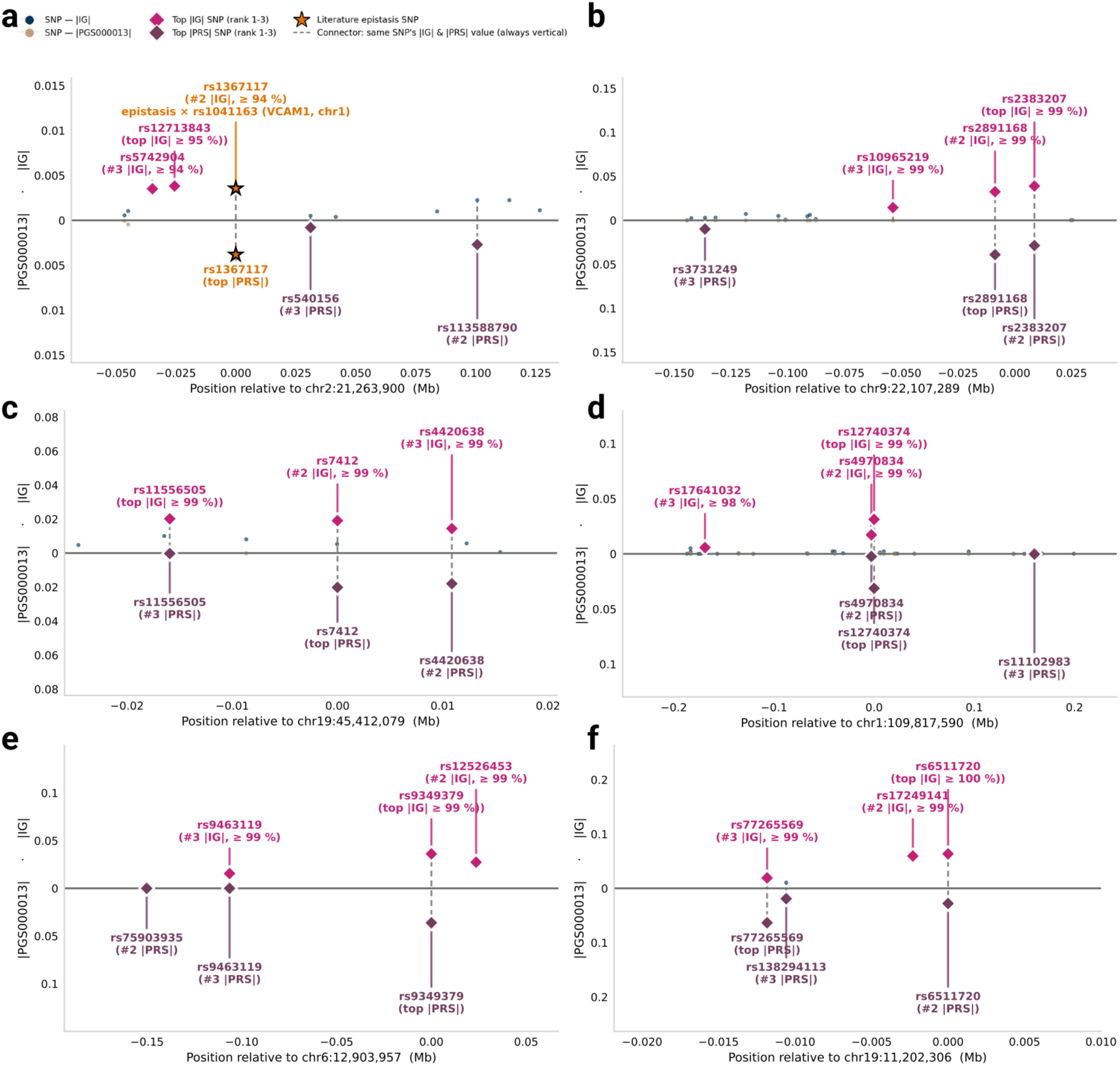
Locus-level convergence between Integrated Gradients and an independent CAD polygenic score at six additional canonical loci. Locus-level Miami plots for six genes identified by inclusion of epistasis variants and joint IG/PGS percentile ranking (Methods) that rank among the most recognizable coronary artery disease and lipid-trait loci in human genetics. (a) APOB; (b) CDKN2B-AS1 (9p21); (c) APOE; (d) CELSR2/SORT1; (e) PHACTR1; (f) LDLR. For each locus, the top 1–3 SNPs by |IG| (magenta) and by |PGS000013| weight (plum) are shown; SNPs ranked top-3 by both metrics appear on both axes, connected by a vertical line joining that SNP’s own |IG| and |PGS| value.

**Supplementary Figure 4.**
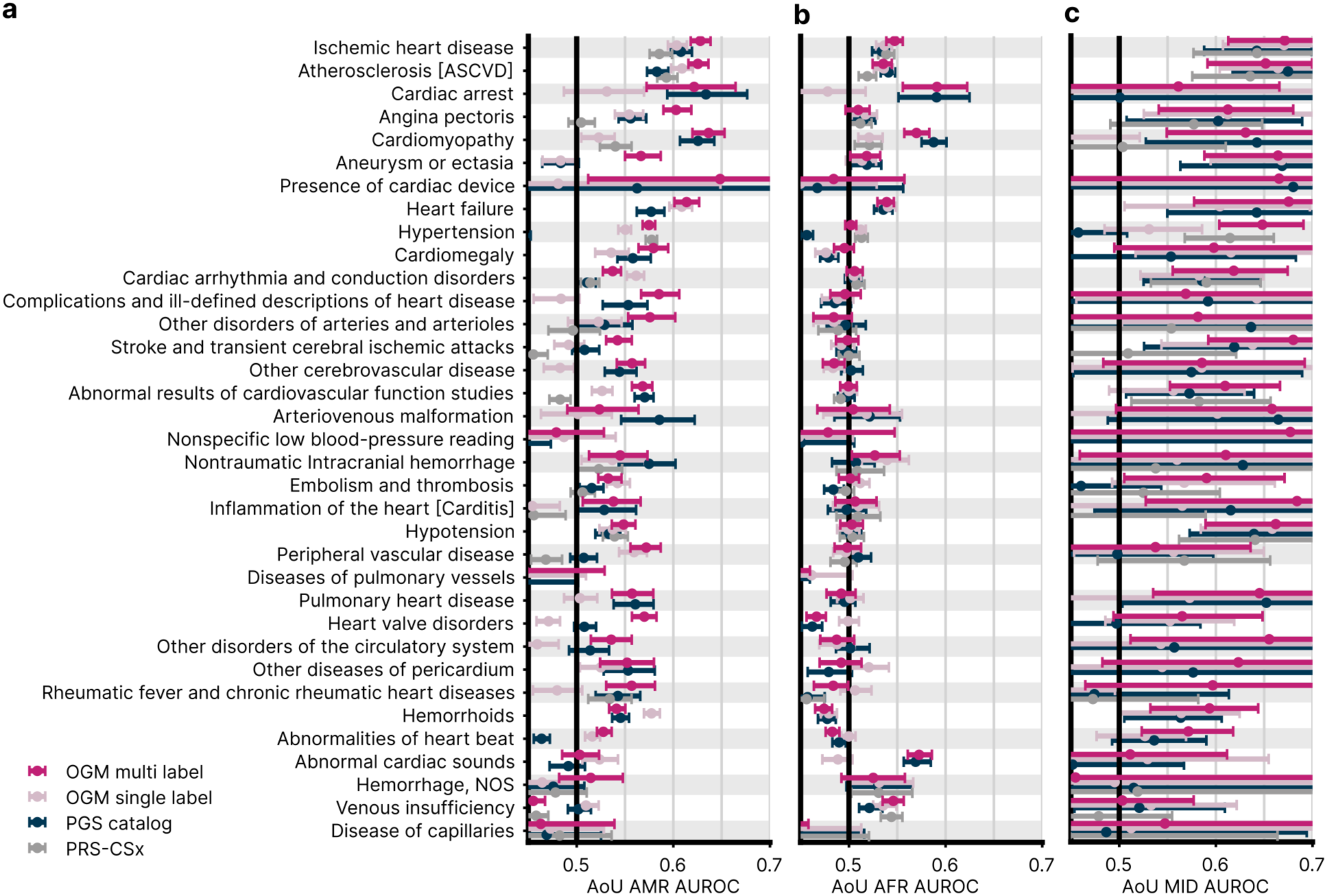
Multilabel predictions for AMR, AFR and MID ancestry groups. All AUROC values are based on genotype, sex and genetic PCs. The multilabel and single-label OGMs are trained in the same set up on UKB data. PRS-CSx models use PanUKBB GWAS. PGS models including covariates are fitted as logistic regressions on the UKB training data used for the OGMs and transferred to AoU. Confidence intervals are 95 percent, based on bootstrapping. Performance for (**a**) AMR, (**b**) AFR, and (**c**) MID populations.

**Supplementary Figure 5.**
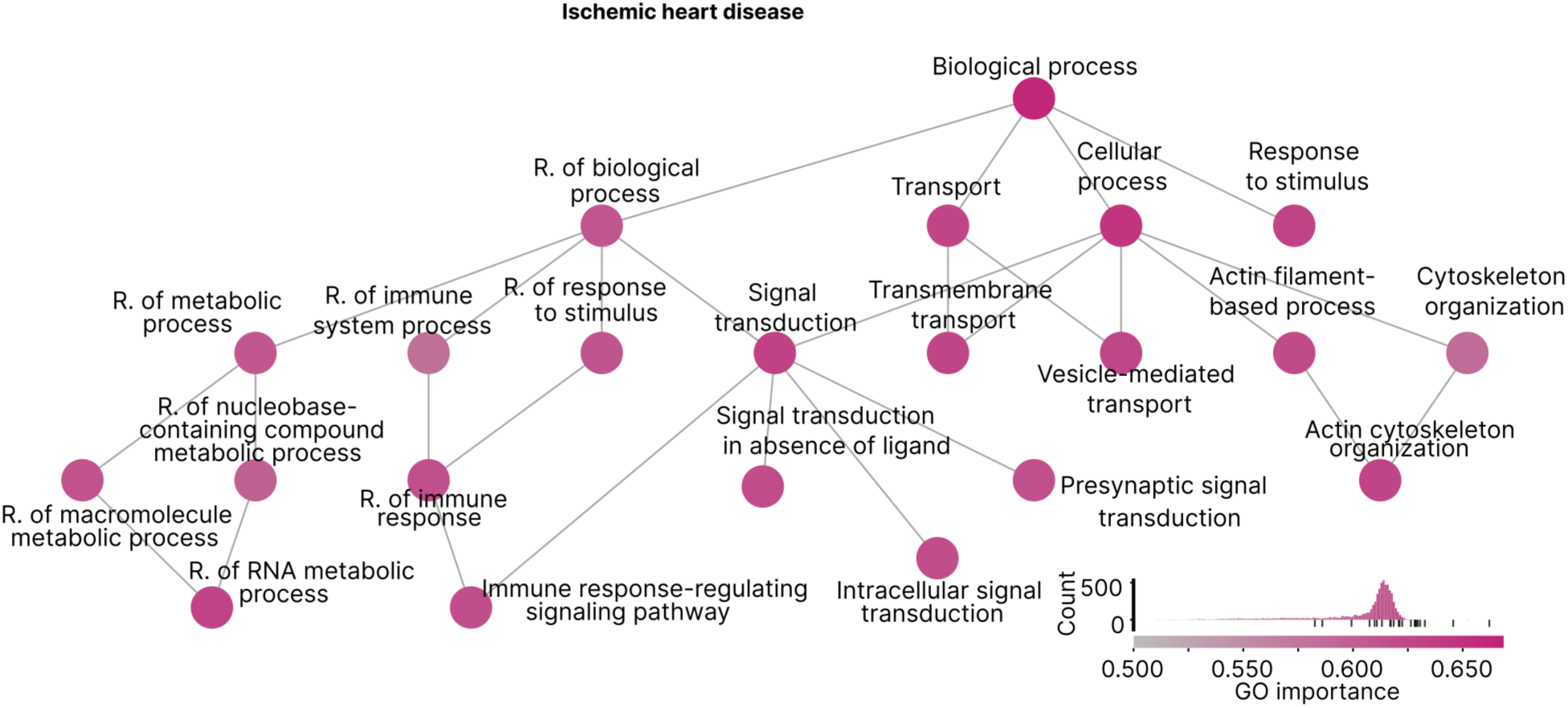
GO importances for ischemic heart disease in a multilabel OGM model. GO importances for the IHD label are shown for the top paths, and top two leaves, including all their parent terms with no manual exclusions. Nodes are coloured by GO importance; the colourscale includes a histogram of all GO importances in the model with black markers indicating the values for the nodes shown in the figure.

**Supplementary Figure 6.**
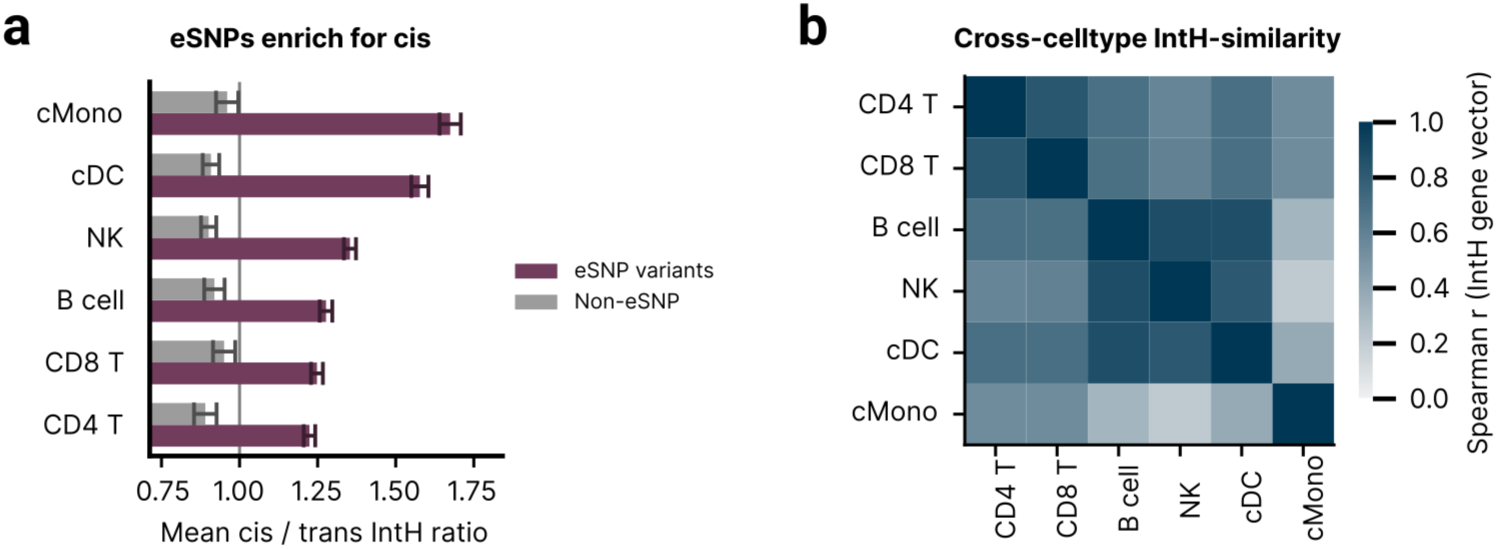
Validation and characterisation of Integrated Hessian-based gene– SNP interaction scores. (**a**) Cis/trans IntH enrichment ratio for eSNP versus non-eSNP anchors, per cell type - eSNP anchors show consistent cis-enrichment (ratio > 1) while non-eSNP anchors do not, recapitulating the cis-dominant architecture of cell-type-specific eQTLs. (**b**) Cross-cell-type Spearman similarity matrix of gene-level mean IntH vectors, showing the degree to which the IntH landscape is shared versus cell-type-specific. (* p < 0.05, ** p < 0.01, *** p < 0.001).

**Supplementary Figure 7.**
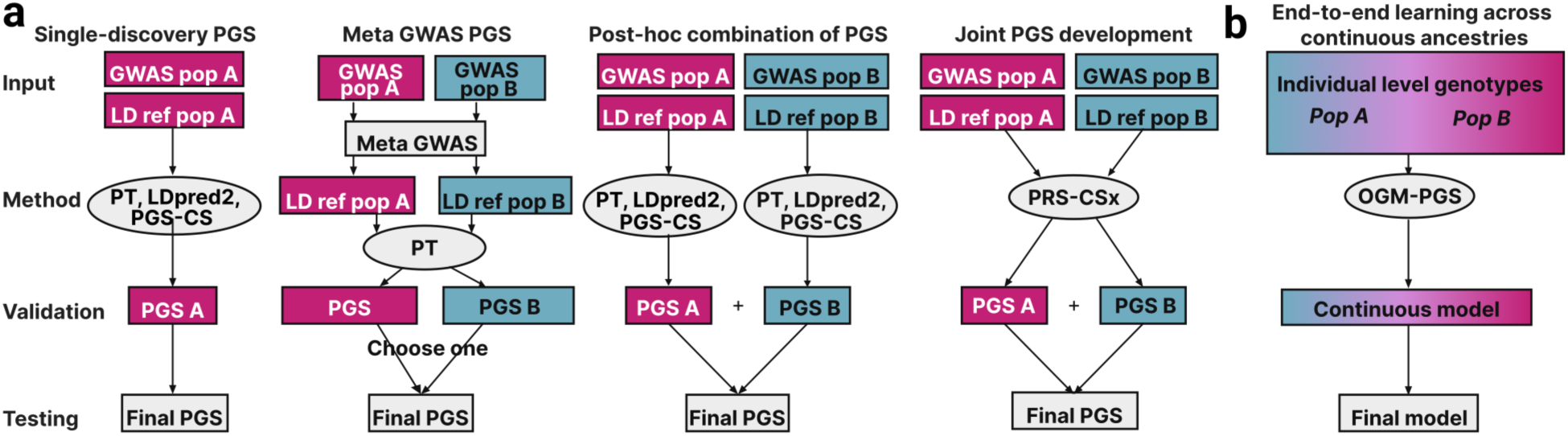
The OGM architecture as a further advance on polygenic prediction methods according to Ruan *et al*. (**a**) Adapted from Figure 1 from Ruan et al.^33^, showing the advances from basic single-discovery PGS methods to joint multi-ancestry methods like PRS-CSx. (**b**) Illustration of the further advance provided by the OGM architecture, namely the continuous representation of ancestry within OGM-based PGS, removing the need for distinct ancestry-assignments.

**Supplementary Table 2.** Model hyperparameter and configurations. Provides the genotypes, phenotypes, and hyperparameters used to train each model.

**[Table in supplementary file]**

**Supplementary Table 3.** Manual mapping of NMR metabolites to ChEBI identifiers. Metabolites with no ChEBI assignment (np.nan) were mapped directly to the Gene Ontology root term “biological process”. Lipoprotein-fraction metabolites (e.g. VLDL_C) carry two ChEBI IDs, one for the chemical class (e.g. CHEBI:16113 for cholesterol) and one for the particle subtype (e.g. CHEBI:39027 for VLDL), linking them to both branches of the ontology hierarchy. Variable names follow the Nightingale Health / UK Biobank NMR metabolomics field naming convention.

**[Table in supplementary file]**

**Supplementary Table 4.** PGS phenotype mappings. Overview of PGS evaluated on the UKB data for cardiovascular phecodes. Columns include the phecodes used in evaluations and comparisons, the PGSCatalog ID, Phecode description, the phenotype description from the PGSCatalog metadata, and the main phenotype description from the PGScatalog. Additionally, UKB performance for the different splits with 95 % confidence intervals is provided for each PGS.

**[Table in supplementary file]**

**Supplementary Table 5.** SLE prediction performance with and without genetic input. Train and test AUROC are shown for the late-fusion model trained with the full frozen OGM genetics latent and with the OGM latent zeroed out during training, for each cell type. Reduced test AUROC under zero-genetics training indicates that the genetic representation contributes non-redundant information and that the model is not merely ignoring it. Inference of the OGM on the genotypes reaches a test AUROC of 0.578.

| Cell type | Test AUROC | Test AUROC (genetics set to 0) |
| --- | --- | --- |
| B cell (b) | 0.666 | 0.634 |
| CD4<sup>+</sup> T (t4) | 0.799 | 0.786 |
| CD8<sup>+</sup> T (t8) | 0.692 | 0.663 |
| Classical monocyte (cm) | 0.835 | 0.829 |
| cDC | 0.780 | 0.754 |
| NK | 0.752 | 0.770 |
| Non-classical monocyte (ncm) | 0.822 | 0.814 |
| pDC | 0.736 | 0.733 |

